# The effectiveness of eight nonpharmaceutical interventions against COVID-19 in 41 countries

**DOI:** 10.1101/2020.05.28.20116129

**Authors:** Jan M. Brauner, Sören Mindermann, Mrinank Sharma, David Johnston, John Salvatier, Tomáš Gavenčiak, Anna B. Stephenson, Gavin Leech, George Altman, Vladimir Mikulik, Alexander John Norman, Joshua Teperowski Monrad, Tamay Besiroglu, Hong Ge, Meghan A. Hartwick, Yee Whye Teh, Leonid Chindelevitch, Yarin Gal, Jan Kulveit

## Abstract

Governments are attempting to control the COVID-19 pandemic with nonpharmaceutical interventions (NPIs). However, it is still largely unknown how effective different NPIs are at reducing transmission. Data-driven studies can estimate the effectiveness of NPIs while minimising assumptions, but existing analyses lack sufficient data and validation to robustly distinguish the effects of individual NPIs. We gather chronological data on NPIs in 41 countries between January and the end of May 2020, creating the largest public NPI dataset collected with independent double entry. We then estimate the effectiveness of 8 NPIs with a Bayesian hierarchical model by linking NPI implementation dates to national case and death counts. The results are supported by extensive empirical validation, including 11 sensitivity analyses with over 200 experimental conditions. We find that closing schools and universities was highly effective; that banning gatherings and closing high-risk businesses was effective, but closing most other businesses had limited further benefit; and that many countries may have been able to reduce R below 1 without issuing a stay-at-home order.

## Introduction

Worldwide, governments have mobilised vast resources to fight the COVID-19 pandemic. A wide range of nonpharmaceutical interventions (NPIs) has been deployed, including drastic measures like stay-at-home orders and the closure of all nonessential businesses. Recent analyses show that these large-scale NPIs were jointly effective at reducing the virus’ effective reproduction number^1^, but it is still largely unknown how effective individual NPIs were. As time progresses and more data become available, we can move beyond estimating the combined effect of a bundle of NPIs and begin to understand the effects of individual interventions. This can help governments efficiently control the epidemic, by focusing on the most effective NPIs to ease the burden put on the population.

A promising way to estimate NPI effectiveness is data-driven, cross-country modelling: inferring effectiveness by relating the NPIs implemented in different countries to the course of the epidemic in these countries. To disentangle the effects of individual NPIs, we need to leverage data from multiple countries with diverse sets of interventions in place. Previous data-driven studies (Table F.8) estimate effectiveness for individual countries^2–4^ or NPIs, although some exceptions exist^1,5–8^ (summarised in Table F.7). In contrast, we evaluate the impact of 8 NPIs on the epidemic’s growth in 34 European and 7 non-European countries. To isolate the effect of individual NPIs, we also require sufficiently diverse data. If all countries implemented the same set of NPIs on the same day, the individual effect of each NPI would be unidentifiable. However, the COVID-19 response was far less coordinated: countries implemented different sets of NPIs, at different times, in different orders (Figure 1).

**Figure 1:**
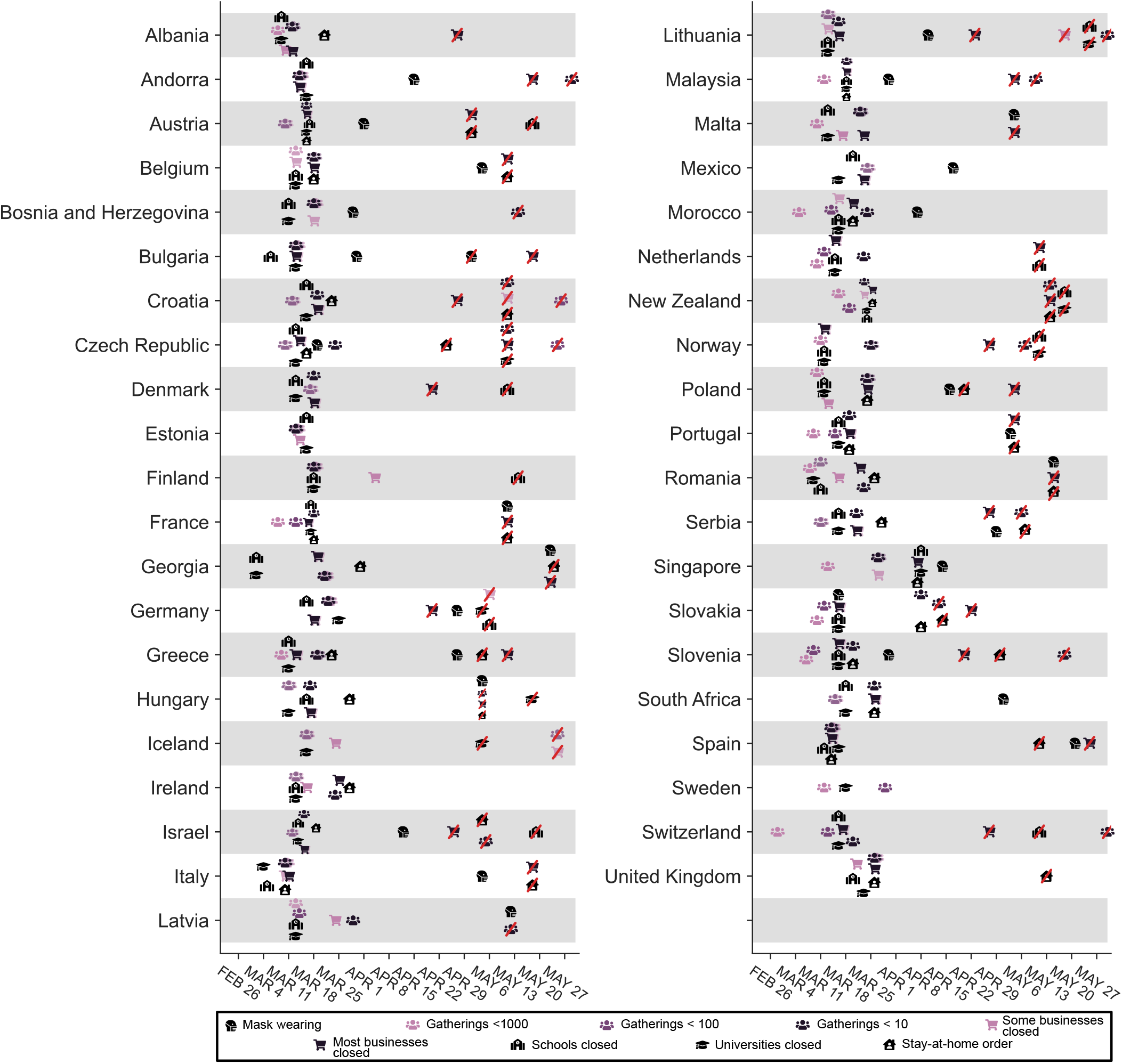
Timing of NPI implementations in early 2020. Crossed-out symbols signify when an NPI was lifted. Detailed definitions of the NPIs are given in Table 1.

Even with diverse data from many countries, estimating NPI effects remains a challenging task. First, models are based on epidemiological parameters that are only known with uncertainty; our NPI effectiveness study, to the best of our knowledge, is the first to include some of this uncertainty in the model. Second, the data are retrospective and observational, meaning that unobserved factors could confound the results. Third, NPI effectiveness estimates can be highly sensitive to arbitrary modelling decisions, as demonstrated by two recent replication studies^9,10^. Fourth, large-scale public NPI datasets suffer from frequent inconsistencies^11^ and missing data^12^. For these reasons, the data and the model must be carefully validated, and insufficiently validated results should not be used to guide policy decisions. We collect the largest public dataset on NPI implementation dates that was validated by independent double entry and perform, to our knowledge, the most extensive validation of any COVID-19 NPI effectiveness estimates to date—a crucial but largely absent or incomplete element of NPI effectiveness studies^10^.

Even with extensive validation, we need to be careful when interpreting this study’s results. We only study the impact NPIs had between January and the end of May 2020, and NPI effectiveness may change over time as circumstances change. In particular, lifting an NPI does not imply that transmission will return to its original level. These and other limitations are detailed in the Discussion.

## Methods

### Dataset

We analyse the effects of NPIs (Table 1) in 41 countries^a^ (see Figure 1). We recorded NPI implementations when the measures were implemented nationally or in most regions of a country (affecting at least three fourths of the population). For each country, the window of analysis starts on the 22nd of January and ends after the first NPI was lifted, or on the 30th of May 2020, whichever was earlier. The reason to end the analysis after the first major reopening^b^ was to avoid a distribution shift. For example, when schools reopened, it was often with safety measures, such as smaller class sizes and distancing rules. It is therefore expected that contact patterns in schools will have been different before school closure compared to after reopening. Modelling this difference explicitly is left for future work. Data on confirmed COVID-19 cases and deaths were taken from the Johns Hopkins CSSE COVID-19 Dataset^13^. The data used in this study, including sources, are available online [here](https://github.com/epidemics/COVIDNPIs/tree/manuscript/online_supplement/data).

**Table 1:** NPIs included in the study. Appendix G details how edge cases in the data collection were handled.

| **NPI** | **Description** |
| --- | --- |
| Mask-wearing mandatory in (some) public spaces | A country has mandated mask usage in the public, sometimes limited to just some public spaces (which the government deems to have a high risk of infection). For example, some countries mandated mask-wearing in most or all indoor public spaces but not outdoors. |
| Gatherings limited to 1000 people or less | A country has set a size limit on gatherings. The limit is at most 1000 people (often less), and gatherings above the maximum size are disallowed. For example, a ban on gatherings of 500 people or more would be classified as “gatherings limited to 1000 or less”, but a ban on gatherings of 2000 people or more would not. |
| Gatherings limited to 100 people or less | A country has set a size limit on gatherings. The limit is at most 100 people (often less). |
| Gatherings limited to 10 people or less | A country has set a size limit on gatherings. The limit is at most 10 people (often less). |
| Some businesses closed | A country has specified a few kinds of customer-facing businesses that are considered “high risk” and need to suspend operations (black-list). Common examples are restaurants, bars, nightclubs, cinemas, and gyms. By default, businesses are not suspended. |
| Most nonessential businesses closed | A country has suspended the operations of many customer-facing businesses. By default, customer-facing businesses are suspended unless they are designated as essential (whitelist). |
| Schools closed | A country has closed most or all schools. |
| Universities closed | A country has closed most or all universities and higher education facilities. |
| Stay-at-home order (with exemptions) | An order for the general public to stay at home has been issued. This is mandatory, not just a recommendation. Exemptions are usually granted for certain purposes (such as shopping, exercise, or going to work), or, more rarely, for certain times of the day. In practice, a stay-at-home order was often accompanied or preceded by other NPIs such as business closures. We account for these other NPIs separately; e.g., when a country issued a law that both closed businesses and ordered the population to stay at home, this is encoded as both a business closure and a stay-at-home order (with exemptions) in the data. In our results, we thus estimate the *additional* effect of the order to generally stay at home. |

#### Data collection

We collected data on the start and end date of NPI implementations, from the start of the pandemic until the 30th of May 2020. Before collecting the data, we experimented with several public NPI datasets, finding that they were not complete enough for our modelling and contained incorrect dates.^c^ By focusing on a smaller set of countries and NPIs than these datasets, we were able to enforce strong quality controls: We used independent double entry and manually compared our data to public datasets for cross-checking.

First, two authors independently researched each country and entered the NPI data into separate spreadsheets. The researchers manually researched the dates using internet searches: there was no automatic component in the data gathering process. The average time spent researching each country per researcher was 1.5 hours.

Second, the researchers independently compared their entries to the following public datasets and, if there were conflicts, visited all primary sources to resolve the conflict: the EFGNPI database^14^, the Oxford COVID-19 Government Response Tracker^15^, and the mask4all dataset^16^.

Third, each country and NPI was again independently entered by one to three paid contractors, who were provided with a detailed description of the NPIs and asked to include primary sources with their data. A researcher then resolved any conflicts between this data and one (but not both) of the spreadsheets.

Finally, the two independent spreadsheets were combined and all conflicts resolved by a researcher. The final dataset contains primary sources (government websites and/or media articles) for each entry.

#### Data Preprocessing

When the case count is small, a large fraction of cases may be imported from other countries and the testing regime may change rapidly. To prevent this from biasing our model, we neglect case numbers before a country has reached 100 confirmed cases and death numbers before a country has reached 10 deaths. We include these thresholds in our sensitivity analysis (Appendix C.3).

### Short model description

In this section, we give a short summary of the model (Figure 2). The detailed model description is given in Appendix A. Code is available online here.

**Figure 2:**
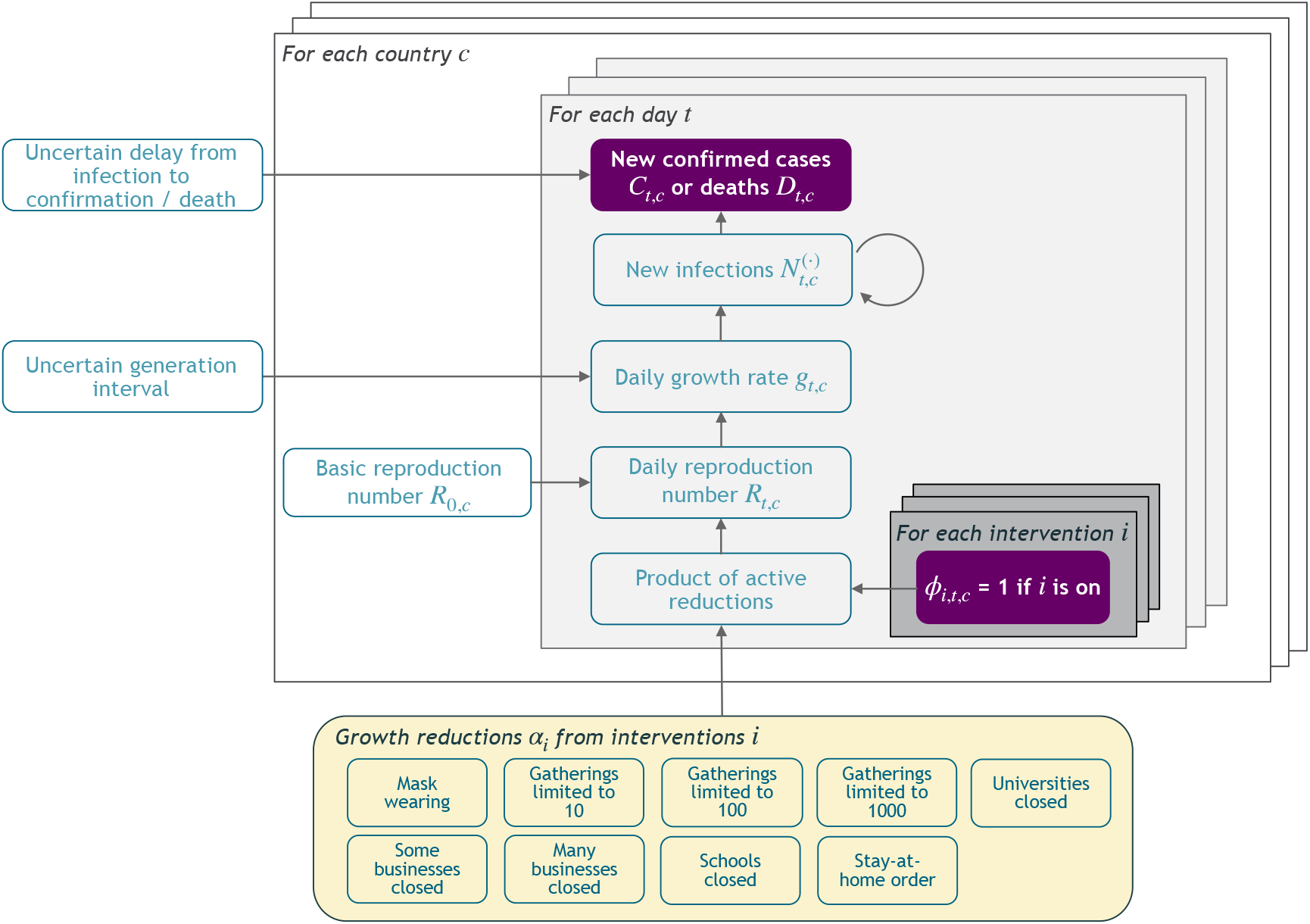
Model Overview. Purple nodes are observed. We describe the diagram from bottom to top: The effectiveness of NPI *i* is represented by *α*_*i*_, which is independent of the country. On each day *t*, a country’s daily reproduction number *R*_*t, c*_ depends on the country’s basic reproduction number *R*_0,*c*_ and the active NPIs. The active NPIs are encoded by F_*i, t, c*_, which is 1 if NPI *i* is active in country *c* at time *t*, and 0 otherwise. *R*_*t, c*_ is transformed into the daily growth rate *g*_*t, c*_ using the generation interval parameters, and subsequently is used to compute the new infections 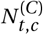 and 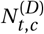 that will turn into confirmed cases and deaths, respectively. Finally, the number of new confirmed cases *C*_*t, c*_ and deaths *D*_*t, c*_ is computed by a discrete convolution of 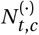 with the respective delay distributions. Our model uses both death and case data: it splits all nodes above the daily growth rate *g*_*t, c*_ into separate branches for deaths and confirmed cases. We account for uncertainty in the generation interval, infection-to-case-confirmation delay and the infection-to-death delay by placing priors over the parameters of these distributions.

Our model uses case and death data from each country to ‘backwards’ infer the number of new infections at each point in time, which is itself used to infer the reproduction numbers. NPI effects are then estimated by relating the daily reproduction numbers to the active NPIs, across all days and countries. This relatively simple, data-driven approach allows us to sidestep assumptions about contact patterns and intensity, infectiousness of different age groups, and so forth, that are typically required in modelling studies. We make several additions to the semi-mechanistic Bayesian hierarchical model of Flaxman et al.^1^, allowing our model to observe *both* cases *and* death data. This increases the amount of data from which we can extract NPI effects, reduces distinct biases in case and death reporting, and reduces the bias from including only countries with many deaths. Since epidemiological parameters are only known with uncertainty, we place priors over them, following recent recommended practice^17^. Additionally, as we do not aim to infer the total number of COVID-19 infections, we can avoid assuming a specific infection fatality rate (IFR) or ascertainment rate (rate of testing).

The growth of the epidemic is determined by the time- and country-specific reproduction number *R*_*t,c*_, which depends on: a) the (unobserved) basic reproduction number *R*_0,*c*_ given no active NPIs and b) the active NPIs at time *t*. *R*_0,*c*_ accounts for all time-invariant factors that affect transmission in country *c*, such as differences in demographics, population density, culture, and health systems^18^. We assume that the effect of each NPI on *R*_*t,c*_ is stable across countries and time. The effectiveness of NPI *i* is represented by a parameter *α*_*i*_, over which we place an Asymmetric Laplace prior that allows for both positive and negative effects but places 80% of its mass on positive effects, reflecting that NPIs are more likely to reduce *R*_*t,c*_ than to increase it. Following Flaxman et al. and others^1,6,8^, each NPI’s effect on *R*_*t,c*_ is assumed to independently affect *R*_*t,c*_ as a multiplicative factor:

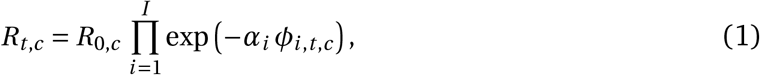

where *ϕ*_*i,c,t*_ *=* 1 indicates that NPI *i* is active in country *c* on day *t* (*ϕ*_*i,c,t*_ *=* 0 otherwise), and *I* is the number of NPIs. The multiplicative effect encodes the plausible assumption that NPIs have a smaller absolute effect when *R*_*t,c*_ is already low. We discuss the meaning of effectiveness estimates given NPI interactions in the Results section.

In the early phase of an epidemic, the number of new daily infections grows exponentially. During exponential growth, there is a one-to-one correspondence between the daily growth rate and *R*_*t,c*_ 19. The correspondence depends on the generation interval (the time between successive infections in a chain of transmission), which we assume to have a Gamma distribution. The prior on the mean generation interval has mean 5.06 days, derived from a meta-analysis^20^.

We model the daily new infection count separately for confirmed cases and deaths, representing those infections which are subsequently reported and those which are subsequently fatal. However, both infection numbers are assumed to grow at the same daily rate in expectation, allowing the use of both data sources to estimate each *α*_*i*_. The infection numbers translate into reported confirmed cases and deaths after a stochastic delay. The delay is the sum of two independent distributions, assumed to be equal across countries: the incubation period and the delay from onset of symptoms to confirmation. We put priors over the means of both distributions, resulting in a prior over the mean infection-to-confirmation delay with a mean of 10.92 days^20^, see Appendix A.3. Similarly, the infection-to-death delay is the sum of the incubation period and the delay from onset of symptoms to death, and the prior over its mean has a mean of 21.8 days^20^. Finally, as in related NPI models^1,6^, both the reported cases and deaths follow a negative binomial noise distribution with an inferred noise dispersion.

Using a Markov chain Monte Carlo (MCMC) sampling algorithm^21^, this model infers posterior distributions of each NPI’s effectiveness while accounting for cross-country variations in testing, reporting, and fatality rates as well as uncertainty in the generation interval and delay distributions. To analyse the extent to which modelling assumptions affect the results, our sensitivity analysis includes all epidemiological parameters, prior distributions, and many of the structural assumptions introduced above (Appendix B.2 and Appendix C). MCMC convergence statistics are given in Appendix C.7.

## Results

### NPI Effectiveness

Our model enables us to estimate the individual effectiveness of each NPI, expressed as a *percentage reduction in R*. As in related work^1,6,8^, this percentage reduction is modelled as constant over countries and time, and independent of the other implemented NPIs. In practice, however, NPI effectiveness may depend on other implemented NPIs and local circumstances. Thus, our effectiveness estimates ought to be interpreted as the *effectiveness averaged over the contexts in which the NPI was implemented, in our data*10. Our results thus give the average NPI effectiveness across typical situations that the NPIs were implemented in. Figure 3 (bottom left) visualises which NPIs typically co-occurred, aiding interpretation.

**Figure 3:**
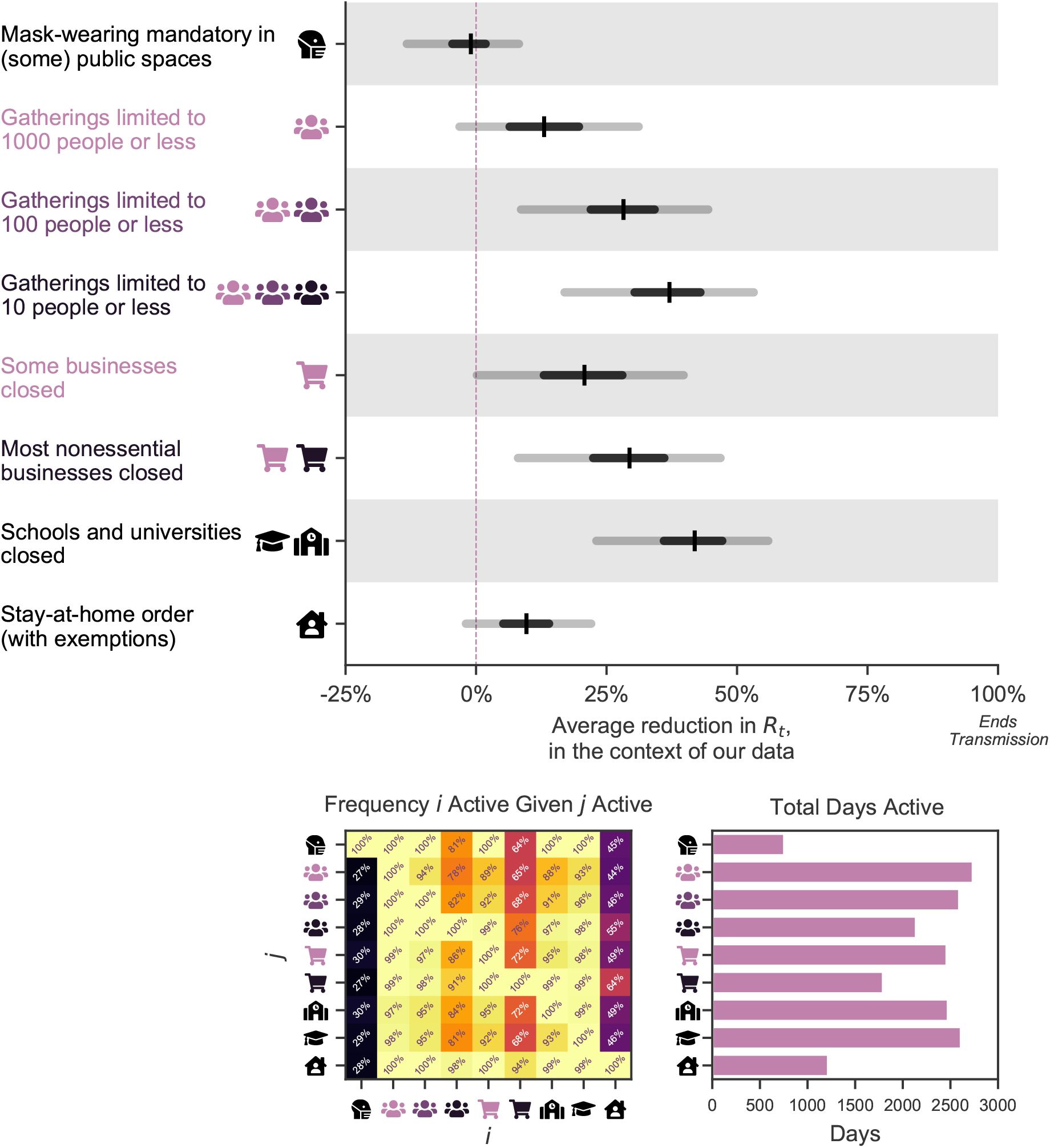
*Top:* NPI effects under default model settings. The Figure shows the average percentage reductions in *R* as observed in our data (or, in terms of the model, the posterior marginal distributions of 1 −exp(−*α*_*i*_)), with median, 50% and 95% credible intervals. A *negative* 1% reduction refers to a 1% increase in *R*. Cumulative effects are shown for hierarchical NPIs (gathering bans and business closures) i.e., the result for *Most nonessential businesses closed* shows the cumulative effect of two NPIs with separate parameters and symbols - closing some (high-risk) businesses, and additionally closing most remaining (non-high-risk, but nonessential) businesses given that some businesses are already closed. *Bottom Left:* Conditional activation matrix. Cell values indicate the frequency that NPI *i* (*x*-axis) was active given that NPI *j* (*y*-axis) was active. E.g., schools were always closed whenever a stay-at-home order was active (bottom row, third column from the right), but not vice versa. *Bottom Right:* Total number of days each NPI was active across all countries.

Under the default model settings, the mean percentage reduction in *R* (with 95% credible interval) associated with each NPI is as follows (Figure 3): mandating mask-wearing in (some) public spaces: −1% (−13%–8%), limiting gatherings to 1000 people or less: 13% (−3%–31%), to 100 people or less: 28% (9%–44%), to 10 people or less: 36% (17%– 53%), closing some high-risk businesses: 20% (0%–40%), closing most nonessential businesses: 29% (8%–47%), closing schools and universities: 41% (23%–56%), and issuing stay-at-home orders (with exemptions): 10% (−2%–22%). Note that we cannot robustly disentangle the individual effects of closing schools and closing universities since the implementation dates of these NPIs coincided nearly perfectly in all countries except Iceland and Sweden (Appendix D.2.1). We thus show the joint effect of closing both schools and universities, and treat “schools and universities closed” as one single NPI going forward.

Some NPIs frequently co-occurred, i.e., were partly *collinear*. However, we are able to isolate the effects of individual NPIs since the collinearity is imperfect and our dataset is large. For every pair of NPIs, we observe one of them without the other for 748 country-days on average (Appendix D.2.2). The minimum number of country-days for any NPI pair is 143 (for limiting gatherings to 1000 or 100 attendees). Additionally, under excessive collinearity, and insufficient data to overcome it, individual effectiveness estimates are highly sensitive to variations in the data and model parameters^22^. High sensitivity prevented Flaxman et al.^1^, who had a smaller dataset, from disentangling NPI effects^9^. Our estimates are substantially less sensitive (see next section). Finally, the posterior correlations between the effectiveness estimates are weak, suggesting manageable collinearity (Appendix D.2.3).

Although the correlations between the individual estimates are weak, we should take them into account when evaluating combined NPI effects. For example, if two NPIs frequently co-occurred, there may be more certainty about the combined effect than about the two individual effects. Figure 4 shows the combined effectiveness of the sets of NPIs that are most common in our data. Together, our set of NPIs reduced *R* by 77% (74%–79%) on average. Across countries, the mean *R* without any NPIs (i.e. the *R*_0_) was 3.3 (Table D.5 reports *R*_0_ for all countries). Starting from this number, the estimated *R* likely could have been reduced below 1 by closing schools and universities, high-risk businesses, and limiting gathering sizes. Readers can interactively explore the effects of sets of NPIs at http://epidemicforecasting.org/calc. A CSV file containing the joint effectiveness of all NPI combinations is available online [here](https://github.com/epidemics/COVIDNPIs/tree/manuscript/online_supplement/results).

**Figure 4:**
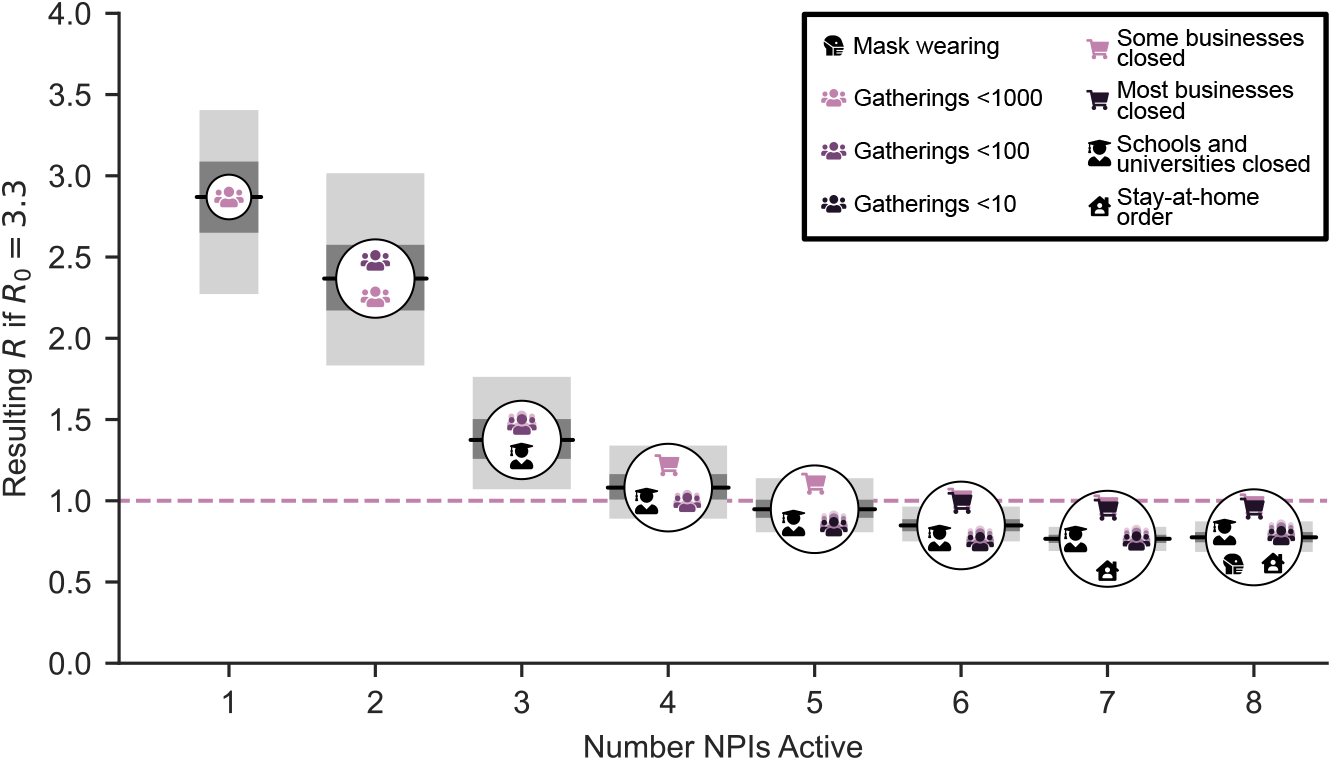
Combined NPI effectiveness for the most common sets of NPIs in our data, by size of the NPI set. Shaded regions denote 50% and 95% credible intervals. *Left*: Maximum *R*_0_ that could be reduced to below 1 for each set of NPIs. *Right*: Predicted *R* after implementation of each set of NPIs, assuming *R*_0_ *=* 3.8. Readers can interactively explore the effects of all sets of NPIs at http://epidemicforecasting.org/calc.

### Sensitivity and validation

We perform a range of validation and sensitivity experiments (Appendix B, with further experiments in Appendix C). First, we analyse how the model extrapolates to unseen countries and find that it makes calibrated forecasts over periods of up to 2 months, with uncertainty increasing over time. Further, we perform multiple sensitivity analyses, studying how results change if we modify the priors over epidemiological parameters, exclude countries from the dataset, use only deaths *or* confirmed cases as observations, vary the data preprocessing, and more. Finally, we investigate our key assumptions by showing results for several alternative models (structural sensitivity^10^) and examine possible confounding of our estimates by unobserved factors influencing *R*. In total, we consider 201 alternative experimental conditions.

Figure 5 (left) shows the median NPI effectiveness across these 201 experimental conditions. Compared to the results under our default settings (Figures 3 and 4), median NPI effects vary under alternative plausible experimental conditions. However, the trends in the results are robust, and some NPIs outperform others under all tested conditions. While we test over large ranges of plausible values, our experiments do not include every possible source of uncertainty and the results might change more substantially under experimental conditions we have not tested.

**Figure 5:**
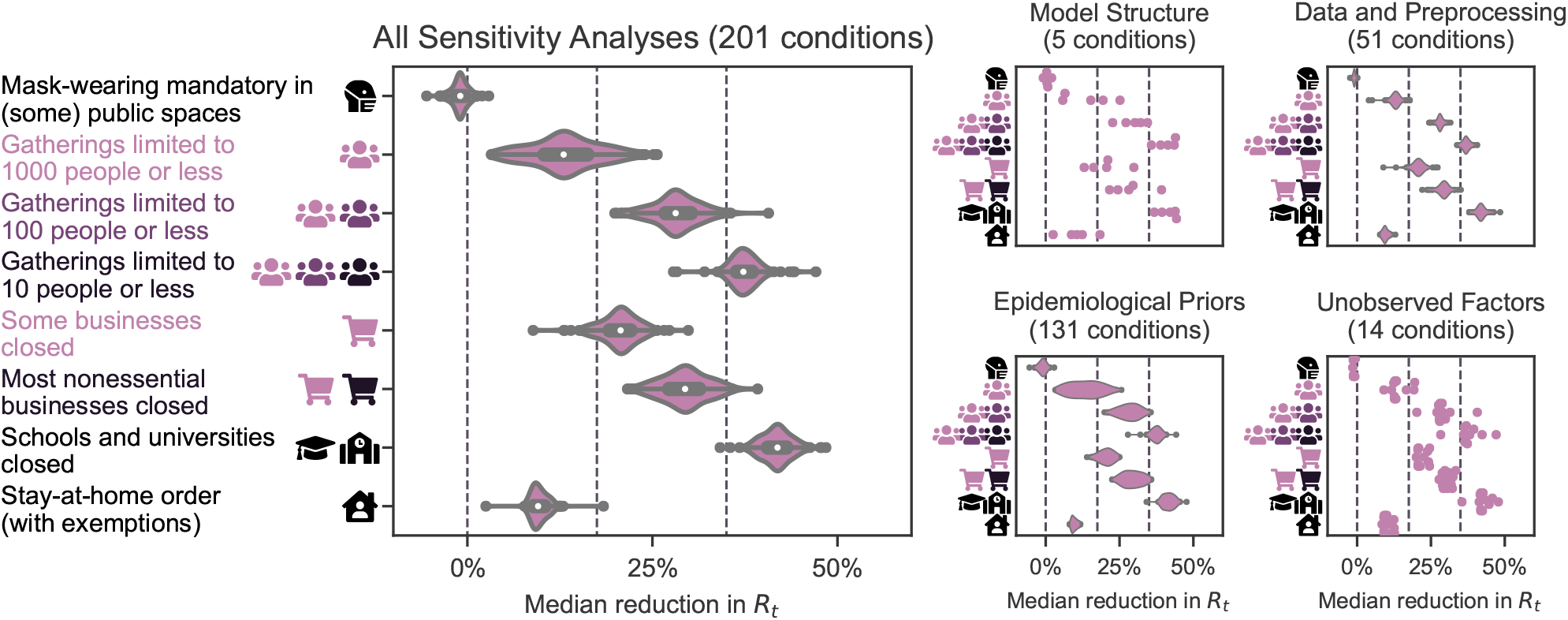
Median NPI effects across the sensitivity analyses. *Left*: Median NPI effects (reduction in *R*) when varying different components of the model or the data in 201 experimental conditions. Results are displayed as violin plots, using kernel density estimation to create the distributions. Inside the violins, the box plots show median and interquartile-range. The vertical lines mark 0%, 17.5%, and 35% (see text). *Right*: Categorised sensitivity analyses. *Structural*: Using only cases or only deaths as observations (2 experimental conditions; Figure B.12), varying the model structure (3 conditions; Figure B.13 left). *Data*: Leaving out one country at a time (41 conditions; Figure B.10), varying the threshold below which cases and deaths are masked (8 conditions; Figure C.17); sensitivity to correcting for undocumented cases and to country-level differences in case ascertainment (2 conditions; Figure B.11). *Epidemiological priors*: *Jointly* varying the means of the priors over the means of the generation interval, the infection-to-case-confirmation delay, and the infection-to-death delay (125 conditions; Figure C.15), varying the prior over *R*_0_ (4 conditions; Figure C.16 left), varying the prior over NPI effectiveness (2 conditions; Figure C.16 right). *Unobserved factors*: Excluding observed NPIs one at a time (9 conditions; Figure B.14 left), controlling for additional NPIs from a different dataset (5 conditions; Figure B.14 right).

We categorise NPI effects into small, moderate, and large, which we define as a median reduction in *R* of less than 17.5%, between 17.5% and 35%, and more than 35% (vertical lines in Figure 5). Six of the NPIs fall into a single category in a large fraction of experimental conditions: school and university closures are associated with a large effect in 99% of experimental conditions, limiting gatherings to 10 people or less in 94%. Closing most nonessential businesses has a moderate effect in 96% of conditions, limiting gatherings to 100 people or less in 97%. Making mask-wearing mandatory in (some) public spaces falls into the “small effect” category in 100% of experimental conditions, issuing stay-at-home orders (with exemptions) in 99%. Two NPIs fall less clearly into one category: Closing some (high-risk) businesses has a moderate effect in 86% of conditions, and limiting gatherings to 1000 people or less has a small effect in 79%. However, both NPIs have small-to-moderate effects in more than 99% of experimental conditions. The effect of limiting gatherings to 1000 people or less is the least stable across the sensitivity analyses, which may reflect its aforementioned partial collinearity with limiting gatherings to 100 people or less.

Aggregating all sensitivity analyses can hide sensitivity to specific assumptions. We display the median NPI effects in four categories of sensitivity analyses (Figure 5, right), and each individual sensitivity analysis is shown in the Appendix. The trends in the results are also stable within the categories.

## Discussion

We use a data-driven approach to estimate the effects that eight nonpharmaceutical interventions had on COVID-19 transmission in 41 countries between January and the end of May 2020. We find that several NPIs were associated with a clear reduction in *R*, in line with the mounting evidence that NPIs can be effective at mitigating and suppressing outbreaks of COVID-19. Furthermore, our results suggest that some NPIs outperformed others. While the exact effectiveness estimates vary with modelling assumptions, the broad conclusions discussed below are largely robust across 201 experimental conditions in 11 sensitivity analyses.

Business closures and gathering bans both seem to have been effective at reducing COVID-19 transmission. Closing only high-risk businesses appears to have been only somewhat less effective than closing most nonessential businesses; the median reduction in *R* differs only by 9% (6%–12%; mean and 95% interval of median estimates across the experiments settings in Figure 5). Closing only high-risk businesses may thus have been the more promising policy option in some circumstances. Limiting gatherings to 10 people or less was more effective than limits up to 100 or 1000 people. This may reflect the fact that small gatherings are common.

As previously discussed, we estimate the average effect each NPI had in the contexts in which it was implemented. When countries introduced stay-at-home orders, they nearly always also banned gatherings and closed schools, universities, and some or most businesses, if they had not done so already (Figure 3, bottom left). Flaxman et al.^1^ and Hsiang et al.^3^ add the effect of these distinct NPIs to the effectiveness of stay-at-home orders, and accordingly find a large effect. In contrast, we account for these other NPIs separately and isolate the *additional* effect of ordering the population to stay at home (when large gatherings are banned and educational institutions and some businesses closed).^d^ In accordance with other studies that took this approach, we find a small effect^2,6^. A typical country could have reduced *R* to below 1 without a stay-at-home order (Figure 4), provided other NPIs were implemented.

Mandating mask-wearing in various public spaces had no clear effect, on average, in the countries we studied. This does not rule out mask-wearing mandates having a larger effect in other contexts. In our data, mask-wearing was only mandated when other NPIs had already reduced public interactions. When most transmission occurs in private spaces, wearing masks in public is expected to be less effective. This might explain why a larger effect was found in studies that included China and South Korea, where mask-wearing was introduced earlier^8,23^. While there is an emerging body of literature indicating that mask-wearing can be effective in reducing transmission, the bulk of evidence comes from healthcare settings^24^. In non-healthcare settings, risk compensation^25^ may play a larger role, potentially reducing effectiveness. While our results cast doubt on reports that mask-wearing is the main determinant shaping a country’s epidemic^23^, the policy still seems promising given all available evidence, due to its comparatively low economic and social costs. Its effectiveness may have increased as other NPIs have been lifted and public interactions have recommenced.

We find a large effect for school and university closures. This finding is remarkably robust across different model structures, variations in the data, and epidemiological assumptions (Figure 5). It remains robust when controlling for NPIs excluded from our study (Figure B.14). Our approach cannot distinguish direct and indirect effects, such as forcing parents to stay at home or causing broader behaviour changes by increasing public concern. Additionally, since school and university closures almost perfectly coincided in the countries we study, our approach cannot distinguish their individual effects (Appendix D.2.1). This limitation likely also holds for other observational studies which do not include data on university closures and estimate only the effect of school closures^1–3,5–8^. Previous evidence on school and university closures is mixed^1,6,26^. Early data suggest that children and young adults are equally susceptible to infection but have a notably lower observed incidence rate than older adults—whether this is due to school and university closures remains unknown^27–30^. Although infected young people are often asymptomatic, they appear to shed similar amounts of virus as older people^31,32^, and might therefore transmit the infection to higher-risk demographics unknowingly. As the role of children in transmission is still unclear^33^ while outbreaks detected in schools are rising^33–35^, this topic merits careful attention.

Our study has several limitations. First, NPI effectiveness may depend on the context of implementation, such as the presence of other NPIs and country-specific factors. Our estimates must be interpreted as the average effectiveness over the contexts in our dataset^10^, and expert judgement is required to adjust them to local circumstances. Second, *R* may have been reduced by unobserved NPIs or spontaneous behaviour changes. To investigate whether these reductions could be falsely attributed to the observed NPIs, we perform several additional analyses and find that our results are stable to a range of unobserved effects (Appendix B.3). However, this sensitivity check cannot provide certainty. Investigating the role of unobserved effects is an important topic to explore further. Third, our results cannot be used without qualification to predict the effect of *lifting* NPIs. For example, closing schools and universities seems to have greatly reduced transmission, but this does not mean that reopening them will cause infections to soar. Educational institutions can implement safety measures such as reduced class sizes as they reopen. Further work is needed to analyse the effects of reopenings; we hope that our collected data aids this effort. Fourth, while we included more NPIs than most previous work (Table F.7), several promising NPIs were excluded. For example, testing, tracing, and case isolation may be an important part of a cost-effective epidemic response^36^, but were not included because it is difficult to obtain comprehensive data. We discuss further limitations in Appendix E.

Although our work focuses on estimating the impact of NPIs on the reproduction number *R*, the ultimate goal of governments may be to reduce the incidence, prevalence, and excess mortality of COVID-19. Controlling *R* is essential, but the contribution of NPIs towards these goals may also be mediated by other factors such as their duration and timing^37^, periodicity and adherence^38,39^, and successful containment^40^. While each of these factors addresses transmission within individual countries, it can be crucial to additionally synchronise NPIs between countries since cases can be imported^41^.

In conclusion, as governments around the world seek to keep *R* below 1 while minimising the social and economic costs of their interventions, we hope that our results can inform policy decisions on which NPIs to implement in any potential further wave of infections. Additionally, our work may provide insight on which areas of public life are most in need of restructuring, so that they can continue despite the pandemic. However, our estimates should not be seen as the final word on NPI effectiveness, but rather as a contribution to a diverse body of evidence, alongside other retrospective studies, simulation studies, experimental trials, and clinical experience.

## Data Availability

All NPI data with sources and model code are available at https://github.com/epidemics/COVIDNPIs/tree/manuscript

## Acknowledgements

Jan Brauner was supported by the EPSRC Centre for Doctoral Training in Autonomous Intelligent Machines and Systems [EP/S024050/1] and by Cancer Research UK. Sören Mindermann’s funding for graduate studies was from Oxford University and DeepMind. Mrinank Sharma was supported by the EPSRC Centre for Doctoral Training in Autonomous Intelligent Machines and Systems [EP/S024050/1]. Gavin Leech was supported by the UKRI Centre for Doctoral Training in Interactive Artificial Intelligence [EP/S022937/1].

The paid contractor work in the data collection and the development of the interactive website was funded by the Berkeley Existential Risk Initiative.

The paid contractor work helping with the data collection, the development of the interactive website, and the costs for cloud compute were funded by the Berkeley Existential Risk Initiative.

## Authors’ contributions

D Johnston, JM Brauner, J Kulveit, G Altman, AJ Norman, JT Monrad, G Leech, V Mikulik designed and conducted the NPI data collection. S Mindermann, M Sharma, JM Brauner, AB Stephenson, H Ge, YW Teh, Y Gal, J Kulveit, T Gavenčiak, J Salvatier, V Mikulik. MA Hartwick, L Chindelevitch designed the model and modelling experiments. M Sharma, AB Stephenson, T Gavenčiak, J Salvatier performed and analysed the modelling experiments. J Kulveit, T Gavenčiak, JM Brauner conceived the research. S Mindermann, M Sharma, JM Brauner, L Chindelevitch, J Kulveit, T Besiroglu did the literature search. JM Brauner, S Mindermann, M Sharma, G Leech, L Chindelevitch, T Besiroglu, V Mikulik wrote the manuscript. All authors read and gave feedback on the manuscript and approved the final manuscript. JM Brauner, S Mindermann, and M Sharma contributed equally. L Chindelevitch, Y Gal and J Kulveit contributed equally to senior authorship.

## Appendix

### Appendix A. Modelling details

#### Appendix A.1. Detailed model description

**Figure A.6:**
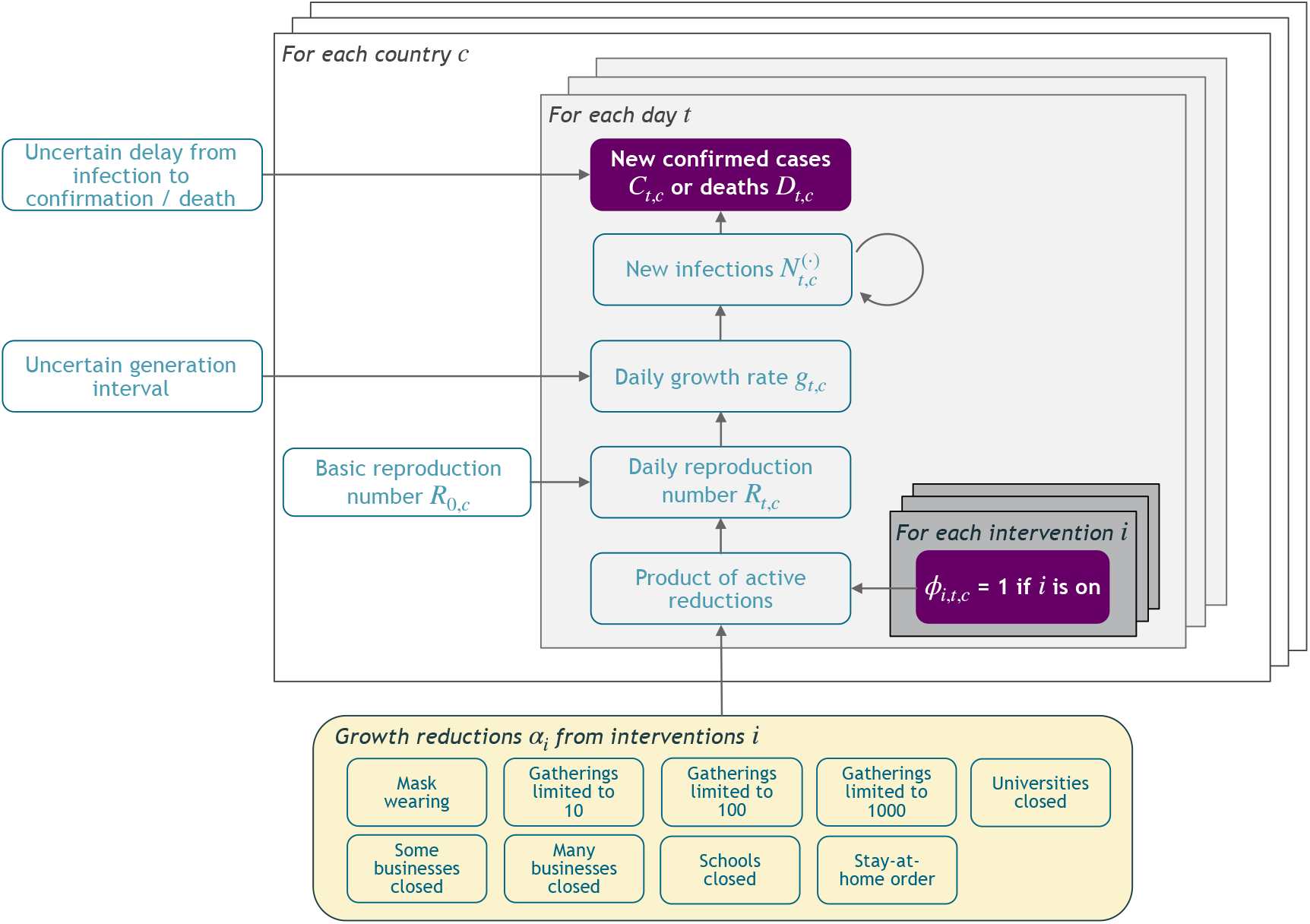
Model Overview. Purple nodes are observed. We describe the diagram from bottom to top: The effectiveness of NPI *i* is represented by *α*_*i*_, which is independent of the country. On each day *t*, a country’s daily reproduction number *R*_*t, c*_ depends on the country’s basic reproduction number *R*_0,*c*_ and the active NPIs. The active NPIs are encoded by F_*i, t, c*_, which is 1 if NPI *i* is active in country *c* at time *t*, and 0 otherwise. *R*_*t, c*_ is transformed into the daily growth rate *g*_*t, c*_ using the generation interval parameters, and subsequently is used to compute the new infections 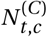 and 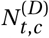 that will turn into confirmed cases and deaths, respectively. Finally, the number of new confirmed cases *C*_*t, c*_ and deaths *D*_*t, c*_ is computed by a discrete convolution of 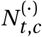 with the respective delay distributions. Our model uses both death and case data: it splits all nodes above the daily growth rate *g*_*t, c*_ into separate branches for deaths and confirmed cases. We account for uncertainty in the generation interval, infection-to-case-confirmation delay and the infection-to-death delay by placing priors over the parameters of these distributions.

We construct a semi-mechanistic Bayesian hierarchical model, similar to Flaxman et al^1^, but with several key extensions. First, we model both confirmed cases *and* deaths, allowing us to leverage significantly more data. Furthermore, we do not assume a specific infection fatality rate (IFR) since we do not aim to infer the *total* number of COVID-19 infections. Additionally, since epidemiological parameters are only known with uncertainty, we place priors over them, following recent best practice^2^. Concretely, we place priors on the means and standard deviations/dispersions of the generation interval, the infectionto-confirmation delay, and the infection-to-death delay. These prior choices are detailed in Appendix A.3. The end of this section details further adaptations which allow us to minimise assumptions about testing, reporting, and the IFR. Code is available online here. For readers who wish to re-implement the model, a concise list of all equations is given in Appendix H.

We describe the model in Figure A.6 from bottom to top. The epidemic’s growth is determined by the time-and-country-specific (instantaneous) reproduction number *R*_*t,c*_. It depends on: a) the basic reproduction number *R*_0,*c*_ without any NPIs active and b) the active NPIs. We place a prior distribution over *R*_0,*c*_, reflecting the wide disagreement of regional estimates of *R*_0_3. The mean *R*_0,*c*_ is 3.28, based on a meta-analysis^4^. We parameterize the effectiveness of NPI *i*, assumed to be same across countries and time, with *α*_*i*_. Each NPI is assumed to have an independent multiplicative effect on *R*_*t,c*_ as follows:

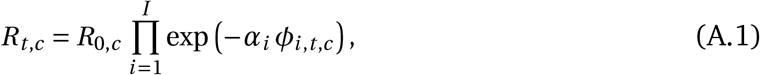

where *ϕ*_*i,c,t*_ *=* 1 means NPI *i* is active in country *c* on day *t* (*ϕ*_*i,c,t*_ *=* 0 otherwise), and *I* is the number of NPIs. We place an Asymmetric Laplace prior over *α*_*i*_ with scale parameter 10, asymmetry parameter 0.5 and location parameter 0. Our prior allows for (unbounded) positive and negative effects as we cannot a priori exclude the possibility that the introduction of an NPI increases *R*. However, our prior places 80% of its mass on positive effects, reflecting a belief that NPIs are much more likely to reduce *R*_*t*_ than not. This is a shrinkage prior, placing 50% of its mass on ‘small’ effectiveness (less than 10% change in *R*_*t*_). All prior distributions are independent.

##### Growth rates

*N*_*t,c*_ denotes the number of new infections at time *t* and country *c*. In the early phase of an epidemic, *N*_*t,c*_ grows exponentially with a daily^a^ growth rate *g*_*t,c*_. During exponential growth, there is a well-known one-to-one correspondence between *g*_*t,c*_ and *R*_*t,c*_ (Eq. 2.9 in^5^):

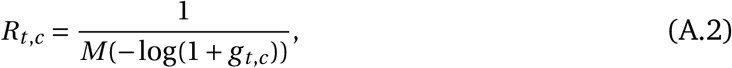

where *M* (·) is the moment-generating function of the distribution of the generation interval (the time between successive cases in a transmission chain). We account for uncertainty in the generation interval (GI), assumed to be a Gamma distribution, by placing prior distributions over its mean and standard deviation (Appendix A.3). Using (A.2), we can write *g*_*t,c*_ as a function *g*_*t,c*_ (*R*_*t,c*_; *µ*_GI_, *σ*_GI_) (see Appendix H), where *µ*_GI_ is the GI mean and *σ*_GI_ is the GI standard deviation.

Note that the GI can change over time due to NPIs. In particular, the effective contact tracing and case isolation in China substantially shortened the mean GI to only 2.6 days among the identified cases^6^. Since most cases were likely not identified even in Wuhan, China^7^, and our study omits most of the countries with highly effective contact tracing programs such as China or South Korea, we model the GI as constant (but uncertain) over time. A possible extension for further work is to model the change in GI explicitly.

##### Infection model

Rather than modelling the total number of new infections *N*_*t,c*_, we model new infections that will either be subsequently a) confirmed positive, 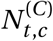, or b) lead to a reported death, 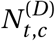. These are inferred from the observation models for cases and deaths. We assume that both grow at the same expected rate, *g*_*t,c*_ :

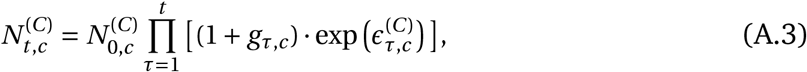

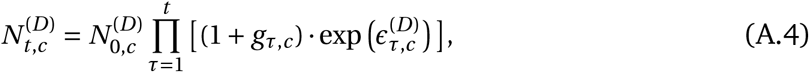

where 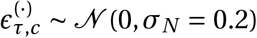 are separate, independent noise terms. Noise on the infection numbers is not used by Flaxman et al.^1^ but has a history in epidemic modelling^8^. Empirically, we find that it leads to substantially more robust effectiveness estimates^9^.

We select *σ*_*N*_ by cross-validation, as no reference is available for it. We evaluate five different values (*σ*_*N*_ ∈ {0.05, 0.1, 0.2, 0.3, 0.4}), with a fixed, randomly chosen validation set of 6 countries. *σ*_*N*_ *=* 0.2 maximises the predictive log-likelihood on the validation set. This ensures a more calibrated model, which is less likely to produce overconfident or unstable estimates^10^. We did not tune any other aspect of the model.

We seed our model with unobserved initial values, 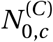 and 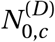, which have uninformative priors.^b^

##### Observation model for confirmed cases

The mean predicted number of new confirmed cases is a discrete convolution

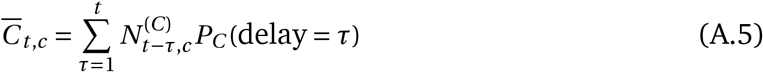

where *P*_*C*_ (delay) is the distribution of the delay from infection to confirmation. In reality, this delay distribution is the sum of two independent distributions: the incubation period (lognormal distribution) and the delay from onset of symptoms to confirmation (negative binomial distribution). These distributions are uncertain but modelling (and placing priors over) them individually leads to unidentifiability. Therefore, we model the *total* delay between infection and confirmation, assuming that this follows a negative binomial distribution. We convert priors over the individual delays to a prior over the total delay by bootstrapping — please see Appendix A.3.

As in Flaxman et al.^1^, the observed cases *C*_*t,c*_ follow a negative binomial noise distribution with mean 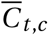 and an inferred dispersion parameter, Ψ^(*C*)^. This distribution encodes that small case numbers are more noisy and should therefore receive less weight. Having separate dispersion parameters for cases and deaths ensures that they can be weighted differently if there is a difference in their noise distributions.

##### Observation model for deaths

The mean predicted number of new deaths is a discrete convolution

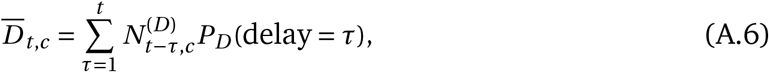

where *P*_*D*_ (delay) is the distribution of the delay from infection to death. As for cases, we model the *total* delay between infection and death as negative binomial, which is in reality the sum of two independent distributions: the incubation period and the delay from onset of symptoms to death (gamma distribution).

Finally, the observed deaths *D*_*t,c*_ also follow a negative binomial distribution with mean 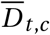 and an inferred dispersion parameter, Ψ^(*D*)^.

The model was implemented in PyMC3^11^. We infer the unobserved variables in our model using the No-U-Turn Sampler (NUTS)^12^, a standard Markov chain Monte Carlo sampling algorithm.

#### Appendix A.2. Testing, reporting, and infection fatality rates

Scaling all values of a time series by a constant does not change its growth rates. The model is therefore invariant to the scale of the observations and consequently to country-level differences in the IFR and the ascertainment rate (the proportion of infected people who are subsequently reported positive). For example, assume countries A and B differ *only* in their ascertainment rates. Then, our model will infer a difference in 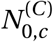 (Eq. (A.3)) but *not* in the growth rates *g*_*t,c*_ across A and B. Accordingly, the inferred NPI effectiveness will be identical.^c^

In reality, a country’s ascertainment rate (and IFR) can also change *over time*. In principle, it is possible to distinguish changes in the ascertainment rate from the NPIs’ effects: decreasing the ascertainment rate decreases future cases *C*_*t,c*_ by a constant factor whereas the introduction of an NPI decreases them by a factor that grows exponentially over time.^d^ The noise term, exp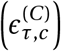 (Eq. (A.3)), mimics changes in the ascertainment rate—noise at time *τ* affects all future cases—and allows for gradual, multiplicative changes in ascertainment.

#### Appendix A.3. Method for uncertainty over key epidemiological parameters

We account for uncertainty in the following delay distributions: the generation interval, the delay from infection to symptom onset (incubation period), the delay from symptom onset to case confirmation, and the delay from symptom onset to death.

Each distribution has an uncertain mean (Table A.3), whose prior distribution we take from a meta-analysis^13^ published shortly after our window of analysis. This helps account for possible heterogeneity between different populations, and therefore often finds higher uncertainty in the means than estimated in primary studies, while using more data. The meta-analysis reports mean values with 95% confidence intervals. We set normal priors over the mean delays, with mean equal to the reported mean in the meta-analysis and standard deviation set such that the prior 95% credible interval includes the 95% confidence intervals from the meta-analysis. For example, the meta-analysis reports an expected mean delay from symptoms to death of 16.71 days with a 95% credible interval ranging from 15.37 to 18.17. We thus assume that the prior is normal with *µ =* 16.71 and *σ =* 0.75, which ensures that its 95% credible interval ranges from 15.25 to 18.17, strictly including the interval reported in the meta analysis to be conservative. Priors are truncated at zero.

Furthermore, we account for the uncertainty in the standard deviation of these delay distributions by placing normal priors based on primary studies, following the same method as above (Table A.4). For the standard deviation of the generation interval, we use patient data from Feretti et al.^14^ and reproduce their method, fitting a Gamma distribution to the generation time.

Finally, the distributional forms of the delay distributions are taken from the above primary studies.

The delay from infection to case confirmation is the sum of the incubation period and the symptom-onset-to-case-confirmation delay. Similarly, the delay from infection to death is the sum of the incubation period and the symptom-onset-to-death delay. However, for computational tractability, we model the *total* delay distributions, converting the priors described above to priors over the total delays. We assume that the total delays follow negative binomial distributions, which are computationally tractable and empirically provide a close fit to both sum distributions. We place normal priors over the mean and dispersion parameters of these total delay distributions by bootstrapping. For example, we compute priors for the total infection-to-case-confirmation delay distribution by sampling *N*_bootstrap_ *=* 250 incubation periods and symptom-onset-to-case-confirmation distributions. For each sampled pair of distributions, we draw 10^6^ samples from their sum and fit a negative binomial using moment matching. We compute the mean and standard deviation of the negative binomial distribution parameters across the bootstrap, and use this for our prior. The resulting priors are given in Table A.2.

**Table A.2:** Epidemiological parameter distributions used in the model. The details on sources and priors used to generate this Table are given in Tables A.3 and A.4.

| Delay type | Sources | Prior on mean | Prior on standard deviation or dispersion | Distributional form |
| --- | --- | --- | --- | --- |
| Infection to case confirmation | Fonfria et al.<sup>13</sup>,<br>Lauer et al.<sup>15</sup>,<br>Cereda et al.<sup>16</sup> | $\mathcal{N}(10.92, \sigma = 0.94)$ | Dispersion:<br>$\mathcal{N}(5.41, \sigma = 0.27)$ | Negative binomial |
| Infection to death | Fonfria et al.<sup>13</sup>,<br>Lauer et al.<sup>15</sup>,<br>Linton et al.<sup>17</sup> | $\mathcal{N}(21.82, \sigma = 1.01)$ | Dispersion:<br>$\mathcal{N}(14.26, \sigma = 5.18)$ | Negative binomial |
| Generation interval | Fonfria et al.<sup>13</sup>,<br>Feretti et al.<sup>14</sup> | $\mathcal{N}(5.06, \sigma = 0.33)$ | SD: $\mathcal{N}(2.11, \sigma = 0.5)$ | Gamma |

**Table A.3:** Mean of epidemiological parameters, all from meta-analysis^13^, and distributional forms.

| Delay type | Reported mean and 95% CI | Prior on mean | Distributional form of delay |
| --- | --- | --- | --- |
| Incubation period | 5.79 (5.48–6.11) | $\log \mu \sim \mathcal{N}(1.53, \sigma = 0.051)$ | Lognormal<sup>15</sup> |
| Onset to death | 16.71 (15.37–18.17) | $\mathcal{N}(16.71, \sigma = 0.75)$ | Gamma<sup>17</sup> |
| Onset to case confirmation | 5.82 (4.74–7.15) | $\mathcal{N}(5.82, \sigma = 0.68)$ | Negative binomial<sup>16</sup> |
| Generation interval | 5.06 (4.50–5.70) | $\mathcal{N}(5.06, \sigma = 0.33)$ | Gamma<sup>14</sup> |

**Table A.4:**
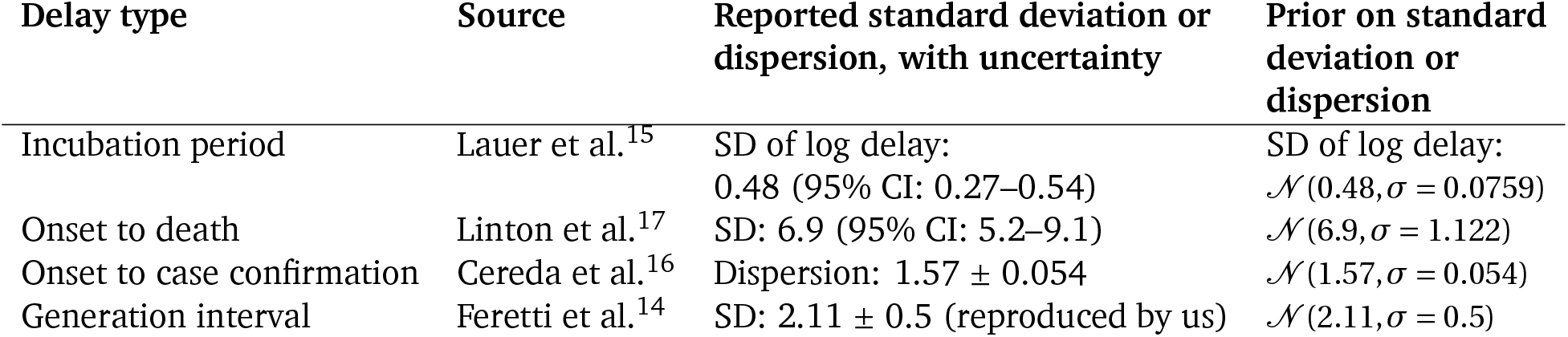
Standard deviation or dispersion of epidemiological parameters.

| Delay type | Source | Reported standard deviation or dispersion, with uncertainty | Prior on standard deviation or dispersion |
| --- | --- | --- | --- |
| Incubation period | Lauer et al.<sup>15</sup> | SD of log delay:<br>0.48 (95% CI: 0.27–0.54) | SD of log delay:<br>$\mathcal{N}(0.48, \sigma = 0.0759)$ |
| Onset to death | Linton et al.<sup>17</sup> | SD: 6.9 (95% CI: 5.2–9.1) | $\mathcal{N}(6.9, \sigma = 1.122)$ |
| Onset to case confirmation | Cereda et al.<sup>16</sup> | Dispersion: $1.57 \pm 0.054$ | $\mathcal{N}(1.57, \sigma = 0.054)$ |
| Generation interval | Feretti et al.<sup>14</sup> | SD: $2.11 \pm 0.5$ (reproduced by us) | $\mathcal{N}(2.11, \sigma = 0.5)$ |

### Appendix B. Validation

#### Appendix B.1. Unseen data

An important way to validate a Bayesian model is by checking its predictions on unseen data, even if prediction is not the purpose of the model^10,18^. If an NPI effectiveness model is entirely unable to extrapolate to unseen countries, we have strong reason to doubt its effectiveness estimates. However, we do not expect NPI effectiveness models to extrapolate perfectly. Almost always, unobserved factors such as changes in the ascertainment rate or IFR, spontaneous behaviour changes, or unobserved NPIs, will affect the observed number of cases and deaths. Our models ought to treat these factors as noise and not attribute their effects on *R*_*t*_ to the observed NPIs.

We use *Leave-One-Out Cross-Validation* (LOO-CV), fitting the model on 40 countries and extrapolating to the excluded country. We repeat this process for all 41 countries. In the excluded country, the first 14 days of case and death data are observed to estimate *R*_0,*c*_ as well as the initial outbreak sizes, 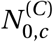 and 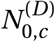. All later days are unobserved (masked). The model uses the effectiveness estimates inferred from the 40 included countries and NPI activation dates to predict the course of the pandemic in the excluded country.

Figure B.7 visually shows extrapolations obtained from LOO-CV for a randomly chosen subset of six countries (all countries are shown in Figures C.18 to C.21). The predictive accuracy of the model, as expected, degrades with an increasing prediction horizon, suggesting the presence of unobserved factors. However, the predictive credible intervals are wide, and increase over time as noise in the growth rate accumulates. Importantly, our model makes calibrated forecasts in excluded countries (Fig. B.8), over periods of up to 2 months.

These are challenging predictions; to the best of our knowledge, no related study analyses how their estimated NPI effects generalise to unseen countries. Most related studies do not validate predictions on unseen data at all^19–23^, reviewed in^9^. Flaxman et al.^1^ hold out the last 14 days, from 20 April to 4th of May, for all countries in aggregate. However, the countries they study implemented all NPIs in March or earlier (Supplementary Table 2 in^1^), meaning that in fact all countries were used to estimate NPI effects.

Note that Fig. B.8 includes the 6 countries that were used to select the hyperparameter (Appendix A). However, the calibration is similar if these 6 countries are excluded (Figure C.23).

#### Appendix B.2. Sensitivity Analysis

Sensitivity analysis reveals the extent to which results depend on uncertain parameters and modelling choices, and can diagnose model misspecification and excessive collinearity in the data^24^. We vary many of the components of our model and recompute the NPI effectiveness estimates, summarised here. Further analysis can be found in Appendix C.

**Figure B.7:**
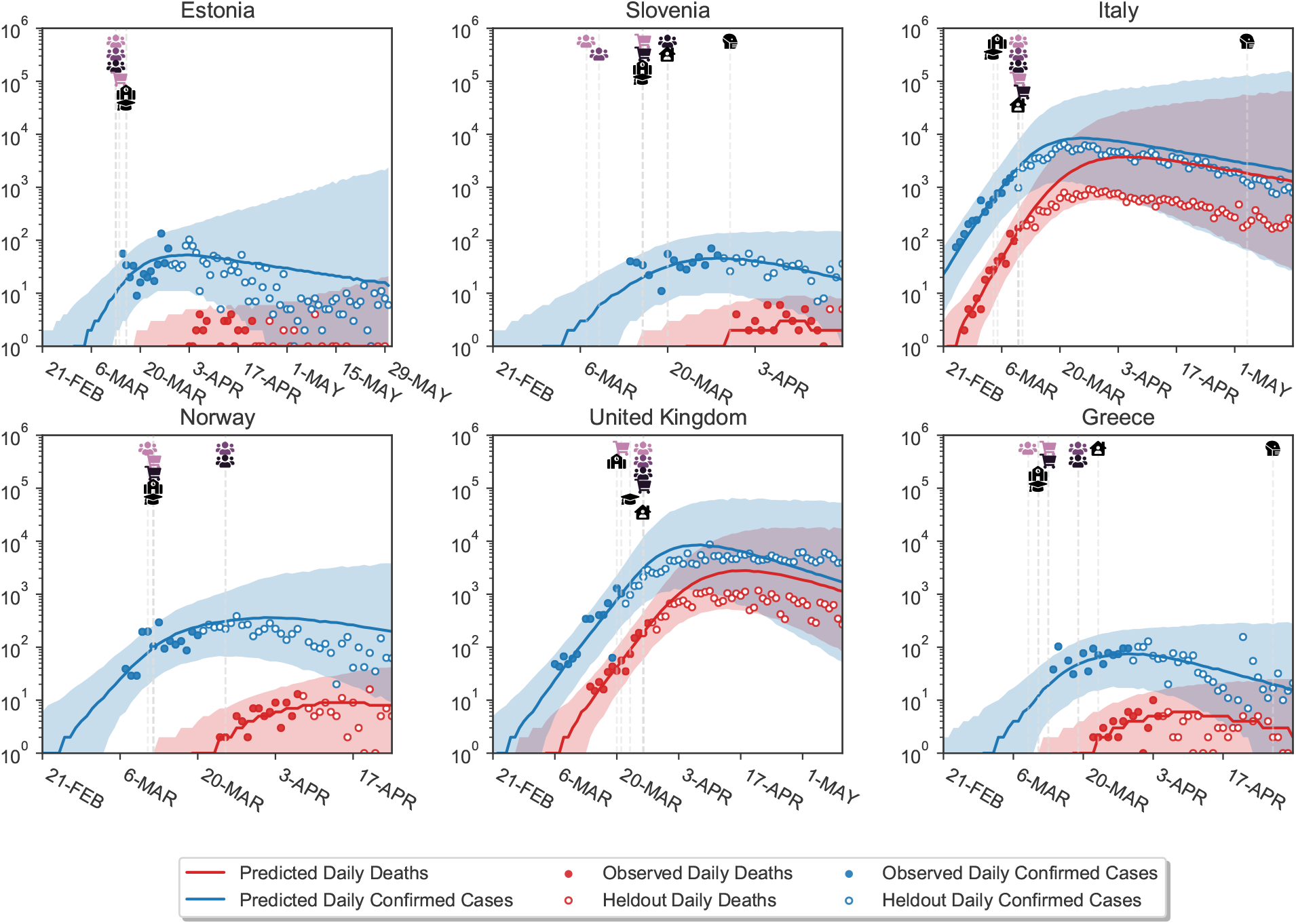
Model extrapolations on a randomly chosen subset of 6 excluded countries, produced using leave-one-out cross-validation. In each excluded country, the first 14 days of death and case data (solid dots) are observed; the model then extrapolates to all future days within the period of analysis (empty dots). For each country, we show the full window of analysis (from the start of the epidemic until the first NPI was lifted, or 30th of May 2020, whichever was earlier; see Methods). The shaded areas represent the 95% credible intervals.

For each sensitivity analysis, we quantify sensitivity using an easily interpretable measure shown above the legend of each plot: the average standard deviation, denoted 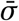. First, for each NPI, we compute the standard deviation of its median effect estimates across all experimental conditions in the given sensitivity analysis. We then average these across the NPIs. Since effectiveness is expressed in percentage reductions of *R*_*t*_, the unit of 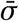 is percent. Zero percent corresponds to no sensitivity.

We perform a multivariate sensitivity analysis on the key epidemiological priors. For computational reasons, all other sensitivity analyses are univariate.

##### Multivariate sensitivity to main epidemiological priors

The key epidemiological parameters in our model describe the delay from infection to case-confirmation, the delay from infection to death, and the generation interval. Our model places prior distributions over the means of these delay distributions. Since the priors already account for uncertainty in the delays, this analysis considers what would happen if our prior beliefs changed. Since the epidemiological parameters may interact in unexpected ways, we perform a multivariate sensitivity analysis, jointly changing the mean of the prior over the mean of each delay distribution (referred to as the “prior mean of the mean” below). We vary the prior means of the mean delays across a wide but epidemiologically plausible range of 5 values, resulting in 125 different experimental conditions. We present these results in two ways.

**Figure B.8:**
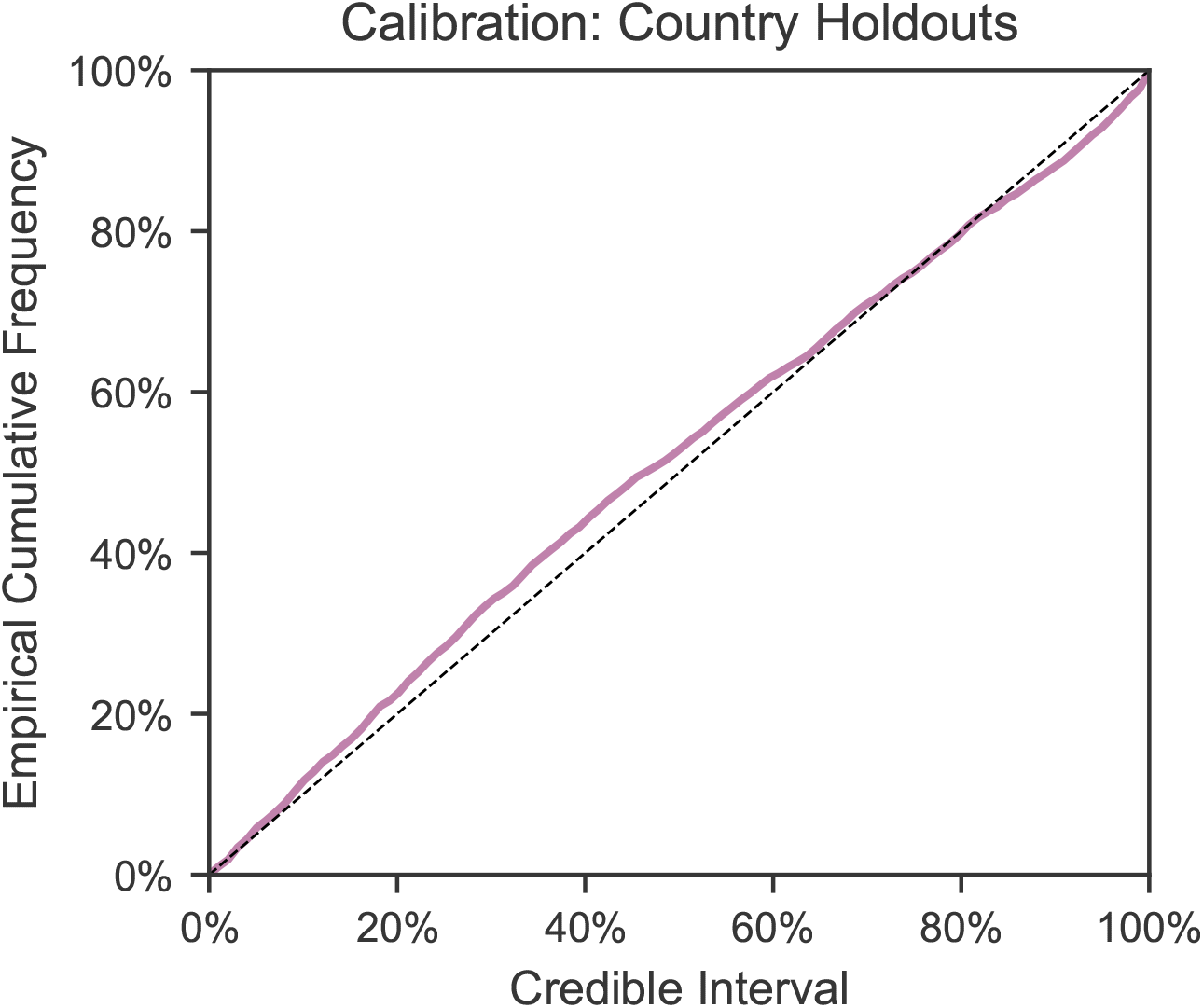
Calibration in held-out countries with leave-one-out cross validation. In each excluded country, the first 14 days of death and case data are observed to estimate *R*_0,*c*_ as well as the initial outbreak sizes; the model then extrapolates to all future days within the period of analysis. The plot shows the percentage of observed daily case and death counts that lie within the X% credible interval, across all countries and days. The identity line represents ideal calibration.

First, Figure B.9 shows the global sensitivity of our results to the prior mean of the mean of each distribution individually. For example, for each setting of the prior mean of the mean generation interval, we show the average over the 5 *×* 5 = 25 runs with different prior means placed over the mean delays between infection and case confirmation/death. This is equivalent to placing a uniform prior across the 125 experiment conditions and marginalising, and is standard methodology for computing global sensitivity to an individual parameter.

The prior means seem to mostly influence the relative effectiveness of business closures and gathering bans. Increasing the prior means of the infection-to-case-confirmation/death mean delays results in larger effectiveness estimates for gatherings bans and smaller estimates for business closures, while increasing the prior mean of the generation interval has the opposite effect.

Second, we assess the sensitivity of our results to *jointly* varying the prior means. This yields 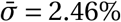. We present this result graphically in Appendix C.1.

**Figure B.9:**
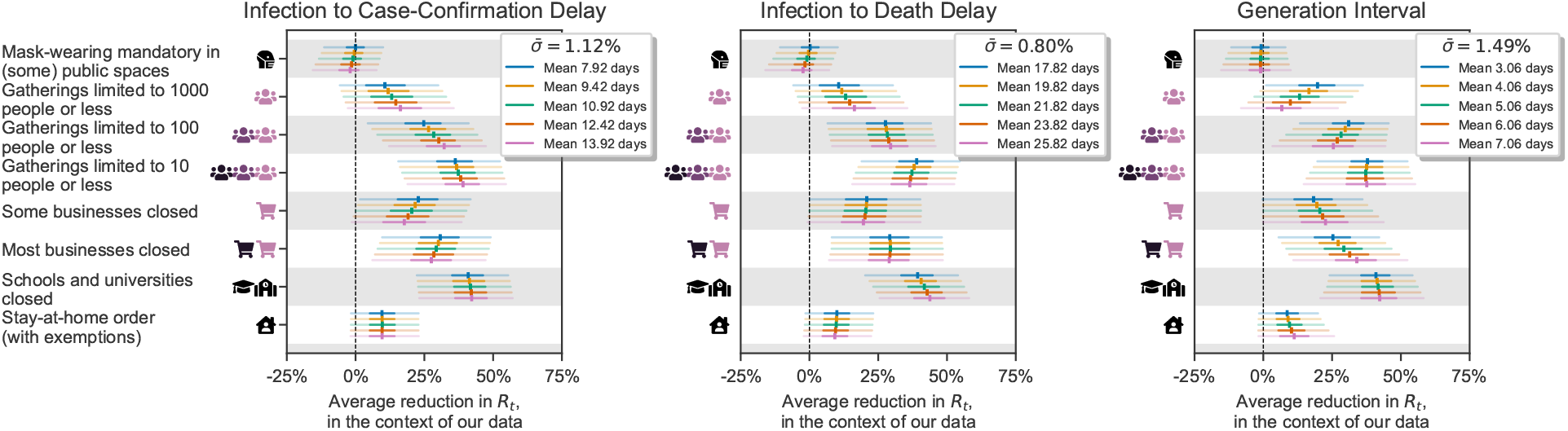
Sensitivity to key epidemiological parameter choices, evaluated using a multivariate analysis. We jointly vary the prior means over the mean of the generation interval, the delay between infection and case confirmation, and the delay between infection and death. Each panel displays sensitivity to the prior mean of one distribution, and the NPI effects are computed by averaging over the 25 runs with different prior means of the two other distributions (see text).

##### Sensitivity to country exclusions

Figure B.10 shows results if one country at a time is excluded from the data. As there is no strong justification for including or excluding any particular country, results ought to be stable if a country is excluded.

**Figure B.10:**
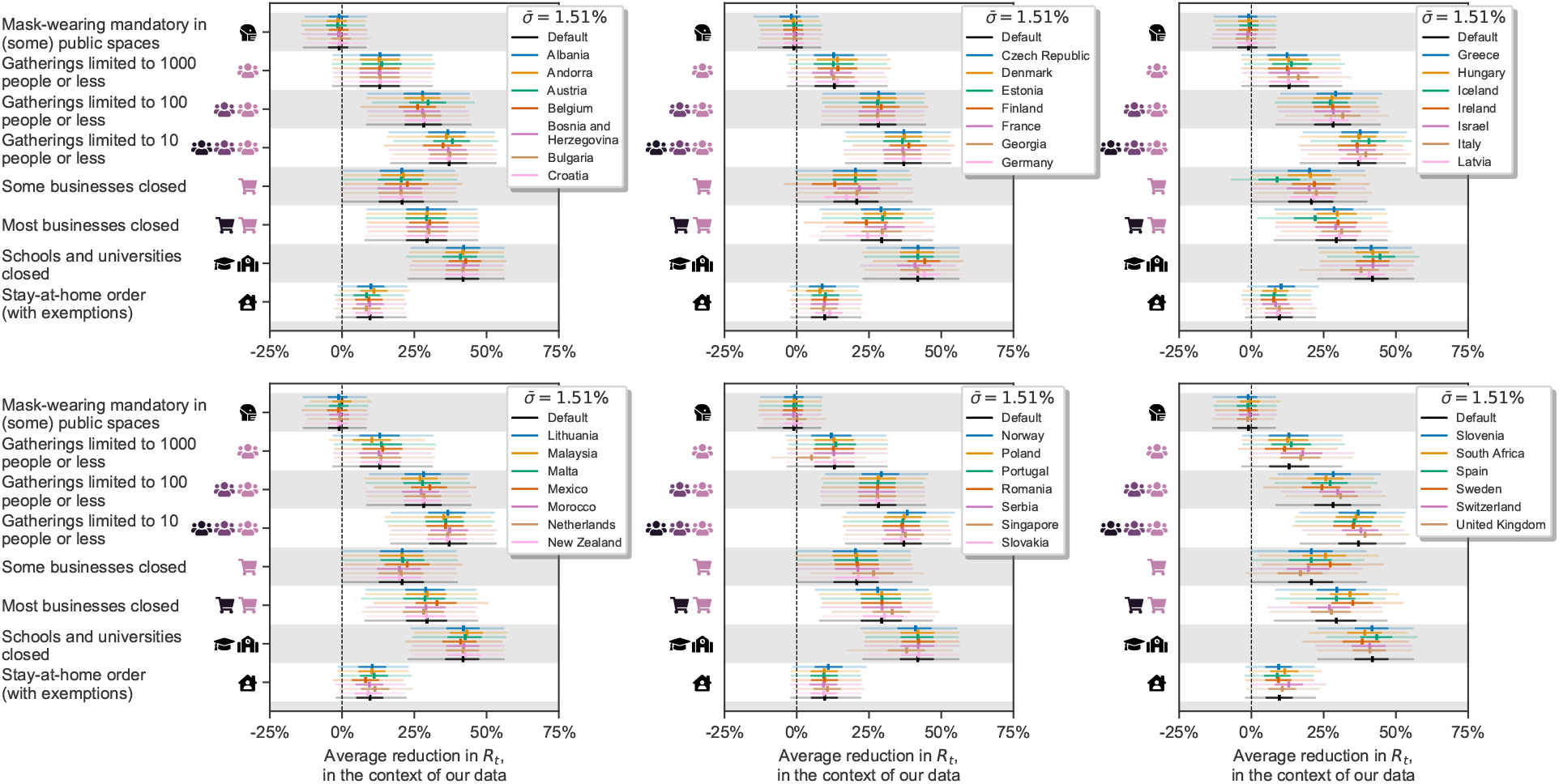
Sensitivity to excluding individual countries.

##### Sensitivity to case-undercounting

Figure B.11 shows results when scaling the recorded number of cases in each country. First, we correct the number of reported cases on each day using estimates of the time-varying ascertainment rate from Golding et al.^25^. For example, if the estimated ascertainment rate in country *c* at time *t* is 50%, we double the corresponding number of reported cases. With this altered case data, the model estimates a larger effect for closing schools and universities, and a smaller effect for limiting gatherings to 1000 people or less. There is little change in the effects estimated for the other NPIs.

Second, we randomly either double or halve the reported number of cases in each country. This is intended to demonstrate that the model is invariant to constant differences in ascertainment between countries. As expected, there are negligible differences compared to the default results.

**Figure B.11:**
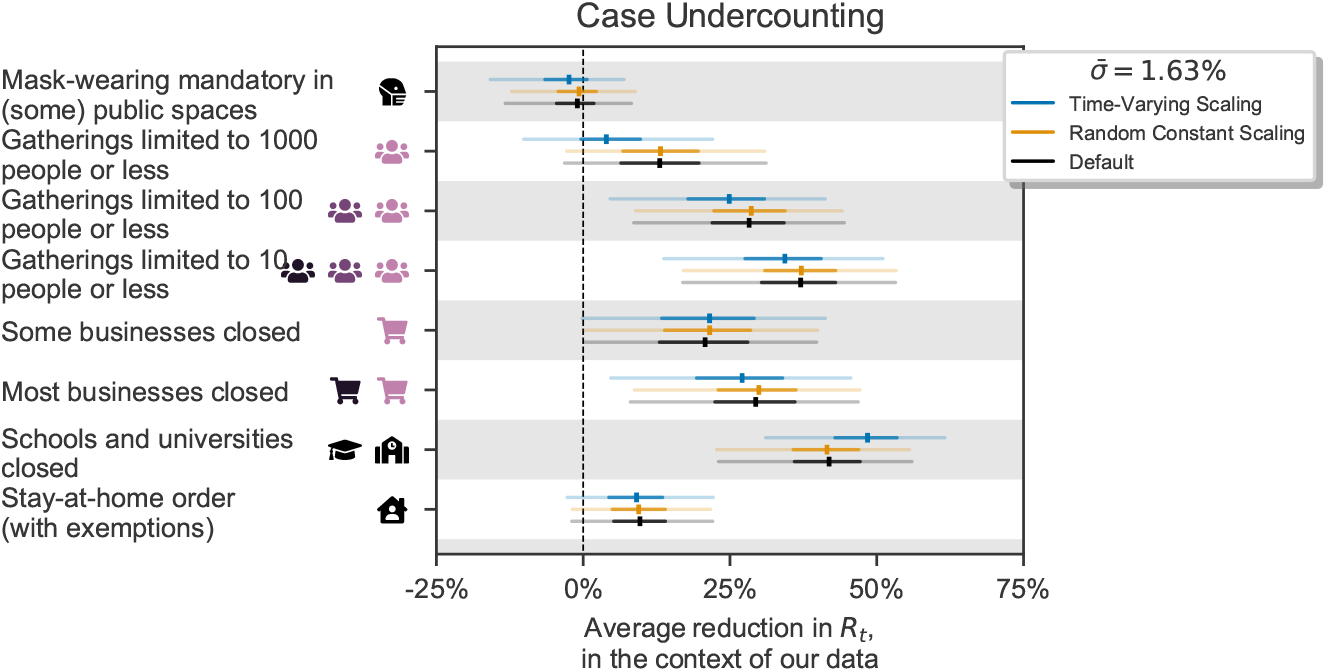
Sensitivity to case undercounting, considering both constant undercounting and time-varying undercounting.

##### Sensitivity to structurally different models

Figure B.12 shows the NPI effectiveness estimates from models that use only cases *or* deaths as observations, in contrast to our main model, which uses both. As expected, there are some differences in the NPI effect estimates between the models, which may be caused by the unique biases present in case data and in death data. Differences could also occur if NPIs differentially affected cases and deaths (e.g. if they affect different age groups). Nevertheless, the study’s qualitative conclusions, as outlined in the Discussion,^e^ hold across the different data types.

A number of implicit structural assumptions are made by the choice of model structure. We test sensitivity to these assumptions by evaluating NPI effectiveness estimates from alternative models, reproducing the structural sensitivity analysis from our concurrent work, where these models are described and compared in detail^9^. While these alternative model structures are also plausible, we chose the model described in the main text as it is relatively simple whilst also being highly robust^9^.

**Figure B.12:**
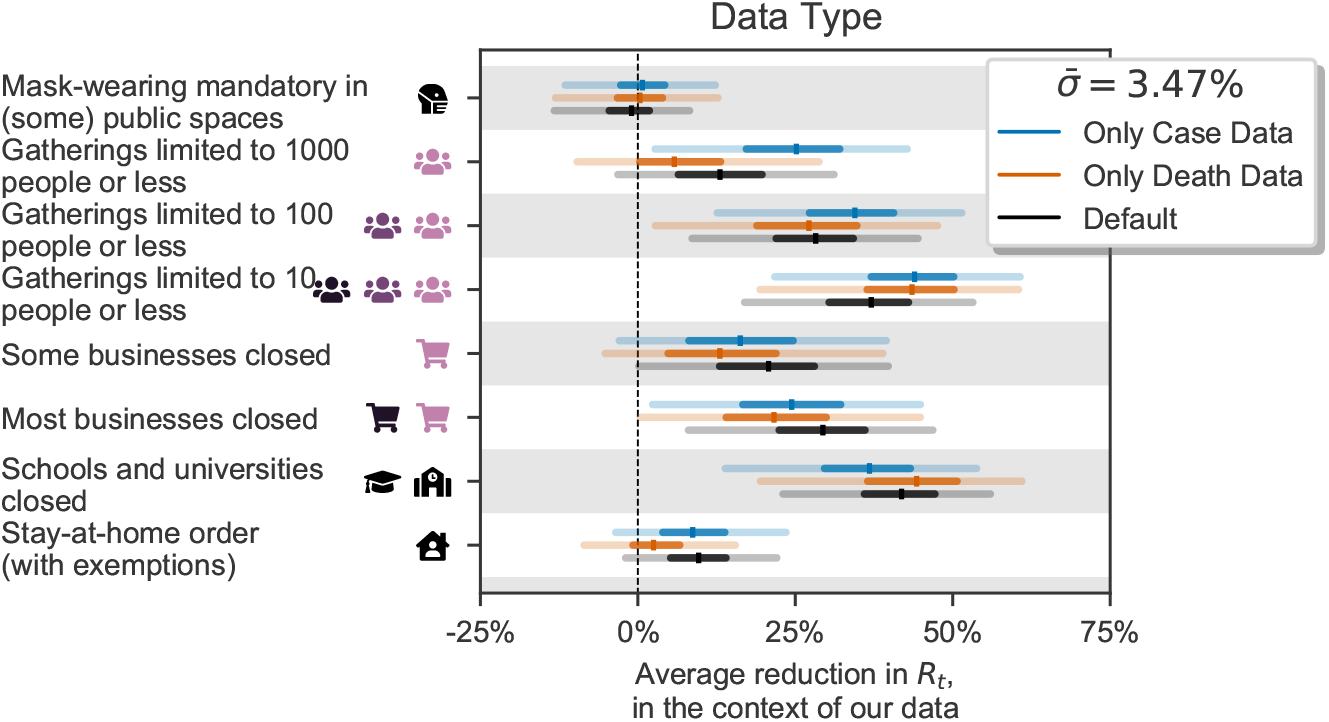
Sensitivity to different data types i.e., models using only confirmed cases, only deaths, or both.

The models are:

1. *Different Effects Model*. The effectiveness of each NPI is allowed to vary across countries.
2. *Discrete Renewal Model*. Instead of converting *R*_*t*_ into a daily growth rate, a renewal process is used as the infection model, as in earlier work^1,8,26–28^. For computational reasons, we do not place a prior distribution over the generation interval parameters for this model.
3. *Noisy-R Model*. The noise terms 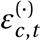 affect *R*_*t*_ rather than the growth rate, as in Fraser^8^.
4. *Additive Effects Model* Each NPI has an additive effect on *R*_*t*_. The joint effectiveness of a set of NPIs is produced by summing, rather than multiplying, their individual effectiveness estimates.

As Figure B.13 (left) shows, the multiplicative effect models support almost all conclusions drawn in the Discussion.^e^ The only exception is that the *stay-at-home order* NPI is categorised as “moderately effective” under the Different Effects Model (median reduction in *R*_*t*_ of 18%).

The results of the Additive Effects Model cannot be directly compared to the other models since it expresses results on a different scale (percentage reduction in *R*_0_ instead of *R*_*t*_). It is therefore shown in a separate panel. However, the trend in the results is visually similar to the default results.

#### Appendix B.3. Robustness to unobserved effects

Our data neither captures all NPIs which were implemented nor directly measures broader behavioural changes. Since these factors influence *R*_*t*_, we must be wary of their effect being attributed to observed NPIs. We investigate this further by assessing how much effectiveness estimates change when previously unobserved factors are included and also when observed factors are excluded. This is best practice for addressing unobserved factors, such as confounders^29,30^.

**Figure B.13:**
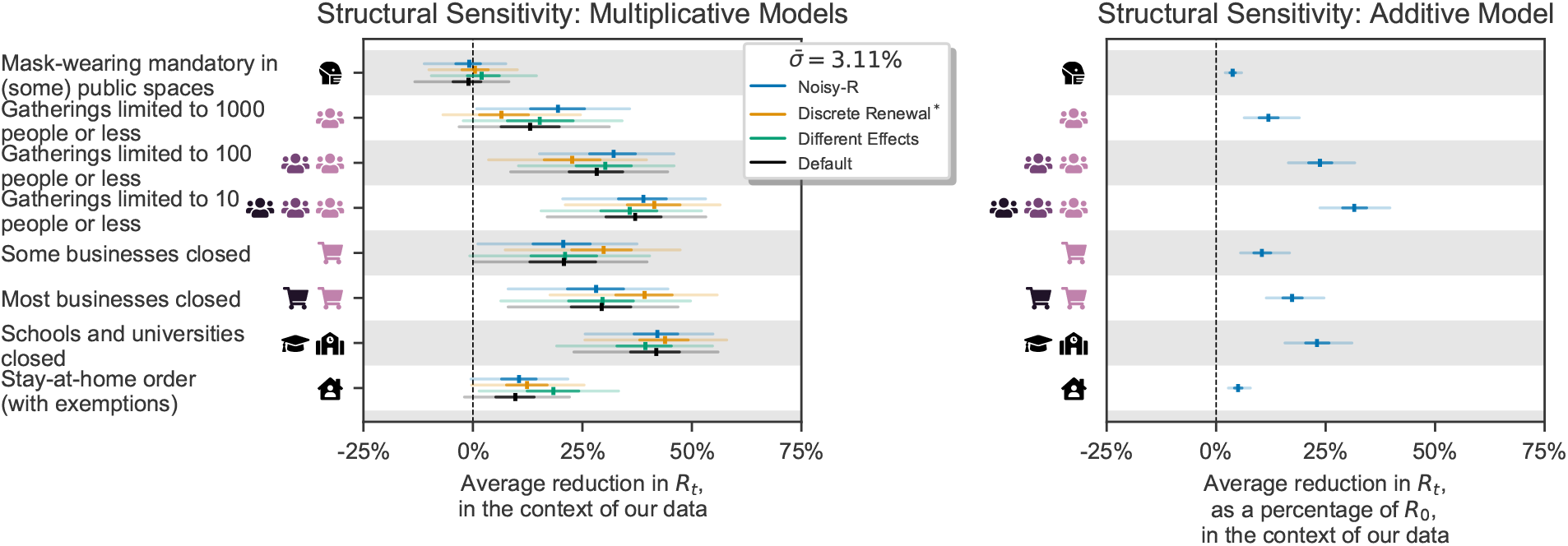
Structural sensitivity analysis. NPI effectiveness estimates under different structural assumptions. *Left*. Effectiveness estimates for multiplicative effect models. Note that, for computational reasons, the discrete renewal model does not have a prior over the generation interval. *Right* Effectiveness estimates for an additive effect models. For this model, the effectiveness of each NPI is expressed as a reduction of *R*_0_ rather than *R*_*t*_.

Unobserved factors can bias results if their timing is correlated with the timing of the observed NPIs^31^. The timings of our *observed* NPIs’ implementation dates are indeed mutually correlated, prompting the question of how much results change when we make previously observed NPIs unobserved. Figure B.14 (left) shows NPI effectiveness estimates when previously observed NPIs are excluded in turn. The study’s main qualitative conclusions^e^ hold across all experimental conditions, and the variations in NPI effectiveness are rarely large enough to cause a change in effectiveness category (Figure 5) except for the *Gatherings limited to 1000 people or less* NPI. Considering that some of the excluded NPIs have strong estimated effects when included, and are correlated with other NPIs, this degree of robustness is encouragingly high. It suggests that unobserved factors will not significantly bias results as long as their effects and their correlations with the studied NPIs do not exceed those of the studied NPIs. We hypothesise that this robustness to unobserved factors is due to the noise on the daily growth rate in our model^9^.

In addition, Figure B.14 (right) shows NPI effectiveness estimates when we observe (i.e. control for) additional NPIs, taken from the OxCGRT dataset^32^.

**Figure B.14:**
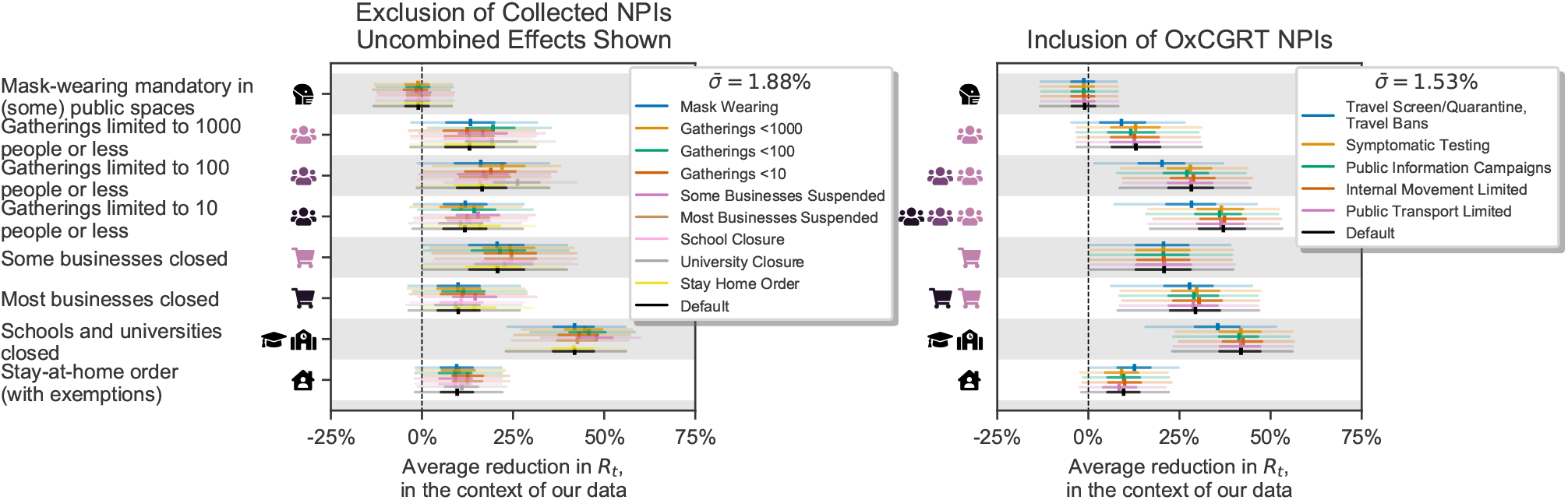
Robustness to unobserved effects. *Left:* Results when excluding previously observed NPIs. We exclude one of the NPIs in turn and show the estimates for the other NPIs. Note that this subplot shows the marginal effect for hierarchical NPIs rather than the total effect, different to other figures that show cumulative effects for gathering bans and businesses closures. For example, Figure 3 displays the total effect of closing most nonessential businesses, while here we show the additional effect of closing most nonessential businesses *over* just closing some high-risk businesses. We show the additional effects here because the effect of a cumulative intervention would become undefined when part of it is excluded from the analysis. *Right:* Results when controlling for previously unobserved NPIs. We include one additional NPI in turn and show the estimates for the NPIs in our study (the additional NPI is not shown).

### Appendix C. Additional sensitivity analyses and validation

#### Appendix C.1. Multivariate Sensitivity Analysis - Expanded Results

We now present the results of our joint multivariate sensitivity analysis (previously summarised in Figure B.9), in which we vary the mean of the prior (“prior mean”) over the mean of the generation interval, the delay between infection and death, and the delay between infection and case confirmation.

Figure C.15 shows, for each NPI, how NPI effects change when all prior means are varied simultaneously.

We find that the most sensitive NPIs are *Gatherings limited to* 1000 *people or fewer, Gatherings limited to* 100 *people or fewer* and *Most nonessential businesses closed*. However, the largest differences appear when all of the parameters are set to the most extreme values that jointly produce a change in a particular direction. We also see systematic trends as the parameters are varied, e.g., increasing the prior mean of the generation interval mean tends to decrease the effectiveness of the *Gatherings limited to 1000 or fewer* NPI, while increasing prior means of the infection to death/case confirmation distribution means tends to increase the effectiveness of this NPI.

**Figure C.15:**
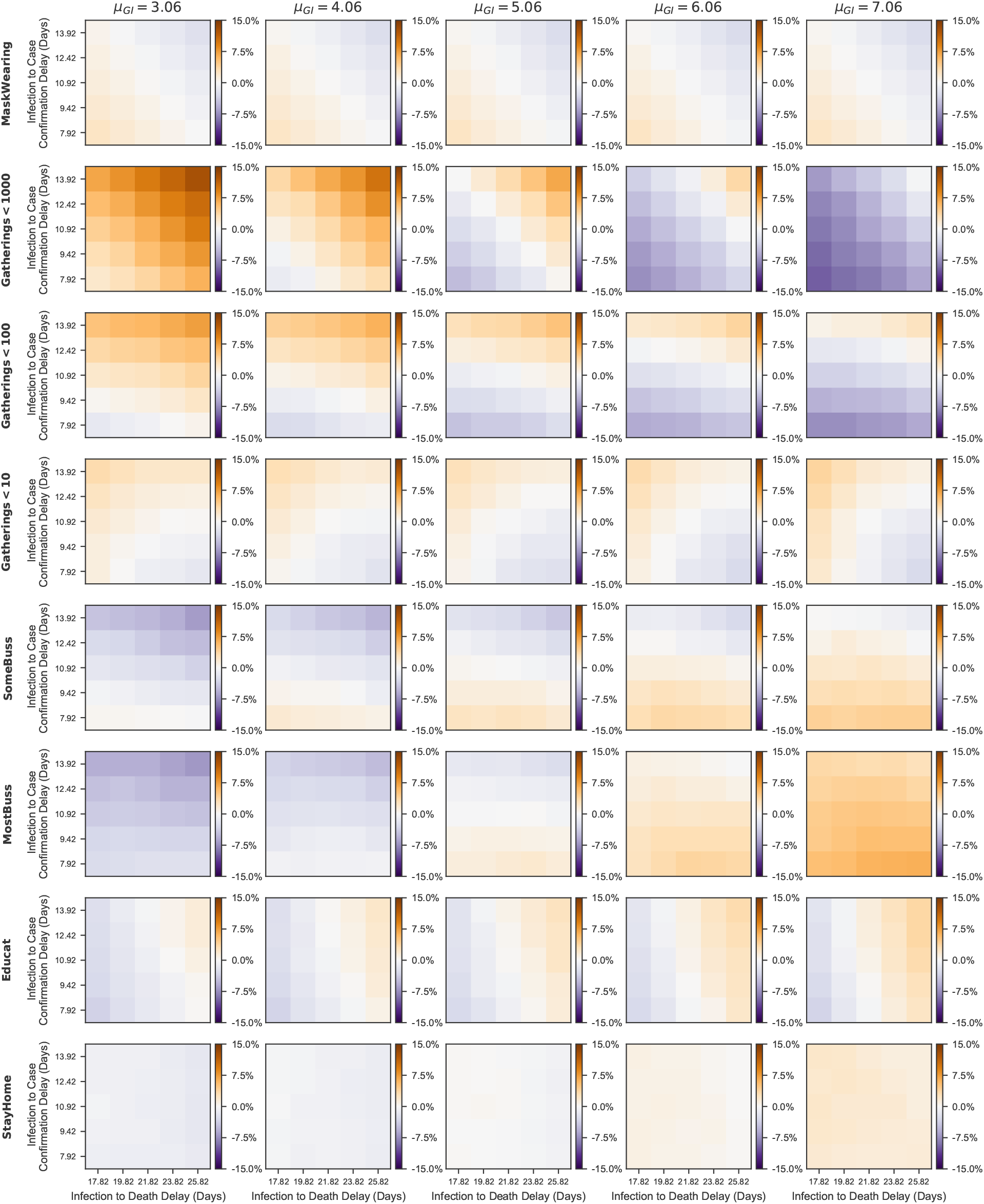
Global sensitivity analysis. Each cell shows the difference between the median effectiveness estimate under a specific experimental condition and the default condition, where the estimates are expressed as percentage reduction in *R*_*t*_. Each subplot represents a particular prior mean of the mean generation interval and an NPI. The NPIs vary top to bottom and the generation interval prior mean increases left to right. Each cell with a subplot represents a particular choice of the prior mean of the mean infection to death delay (increasing left to right) and the prior mean of the mean infection to case confirmation delay (increasing bottom to top). Positive cell values indicate that the median NPI estimate is larger than with default settings and vice versa.

#### Appendix C.2. Sensitivity to additional epidemiological assumptions

For completeness, Figure C.16 shows sensitivity to two further epidemiological assumptions. On the left, we show the dependence of NPI effectiveness estimates on *R*_0_. We place a Normal prior over *R*_0_, and a hyperprior over its variance, reflecting the wide disagreement of regional estimates of *R*_0_. In the sensitivity analysis, we vary the mean of the prior from a low value (2.38) to a high one (4.28). As expected, higher mean *R*_0_ values result in larger NPI effects. This trend is apparent in all NPIs but stronger for NPIs that tended to be implemented early on (like school closures and gathering bans).

On the right, we vary the prior over NPI effectiveness. Our default prior on NPI effectiveness is assymmetric, reflecting a belief that NPIs are more likely to reduce *R*_*t*_ than increase it. Here, we additionally test a symmetric Normal prior, and a one-sided Half-Normal prior, which does not allow for NPIs to increase *R*_*t*_.

#### Appendix C.3. Sensitivity to data preprocessing

When the case count is small, a large fraction of cases may be imported from other countries and the testing regime may change rapidly. To prevent this from biasing our model, we neglect case numbers before a country has reached 100 confirmed cases. Similarly, we neglect death numbers before a country has reached 10 deaths. Here, we vary these thresholds. Intuitively, the minimum cases threshold should be higher than the minimum deaths threshold.

**Figure C.16:**
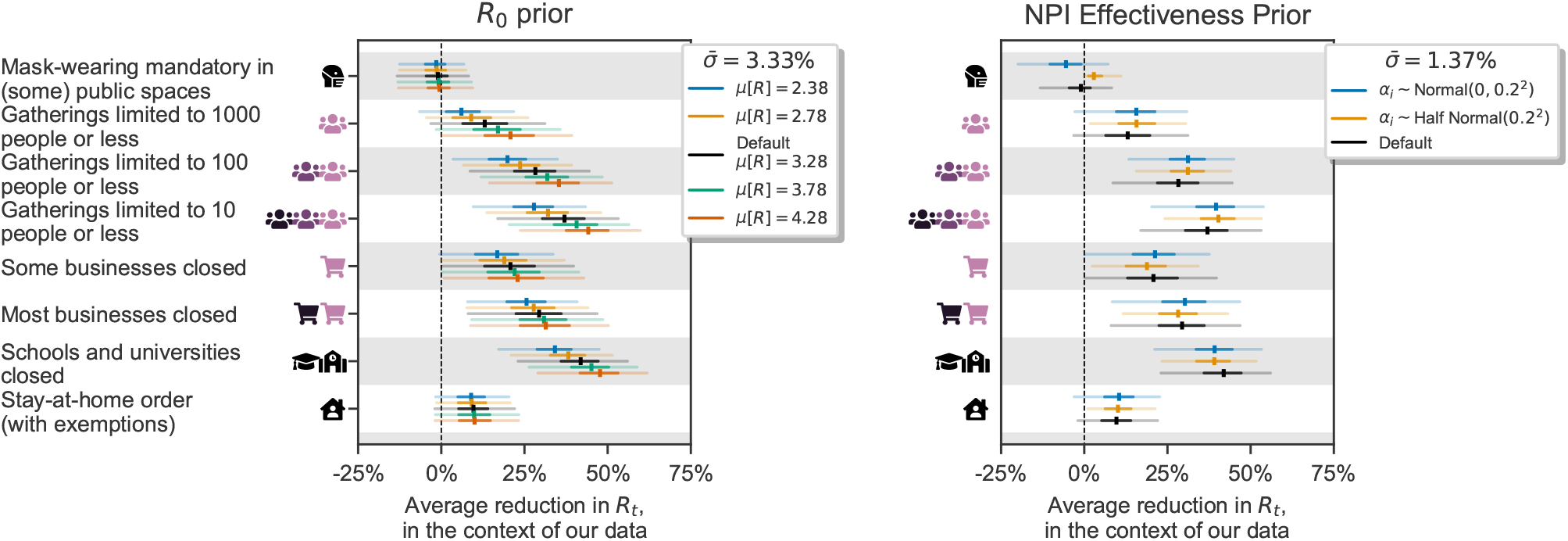
Sensitivity to additional epidemiological priors. *Left:* Sensitivity to the prior mean on *R*_0_. *Right:* Sensitivity to the NPI effectiveness prior.

**Figure C.17:**
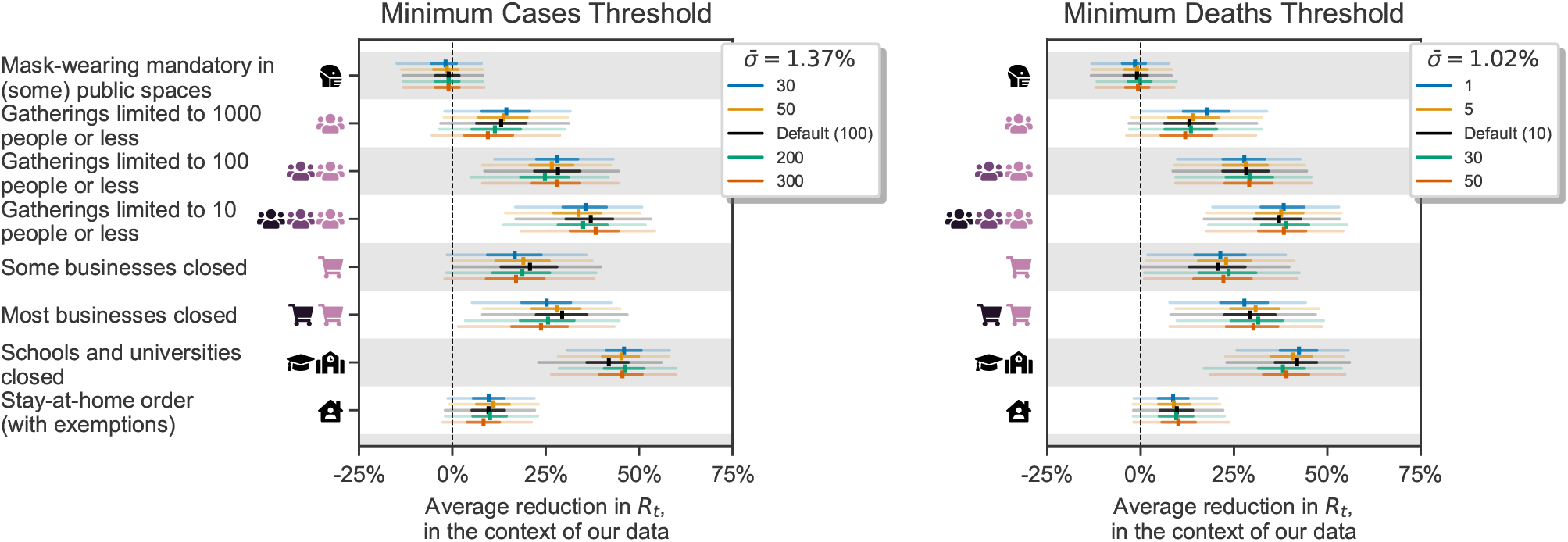
Data preprocessing sensitivity. *Left*. Variations in the minimum number of cases. *Right*. Variations in the minimum number of deaths.

#### Appendix C.4. Validation by predicting unseen data - all countries

Here we show the country-level plots of the single-country holdout experiments from Appendix B.1. We use leave-one-out cross-validation: fitting the model on 40 countries and showing its predictions on the excluded country. We repeat this process for all 41 countries. In the excluded country, the first 14 days (filled dots) of death and case data are observed to allow roughly inferring *R*_0_. These days are not used to infer NPI effectiveness. All later days are masked (unfilled dots). The activation dates of NPIs are also given, and the model uses the effectiveness estimates inferred from the 40 other countries.

**Figure C.18:**
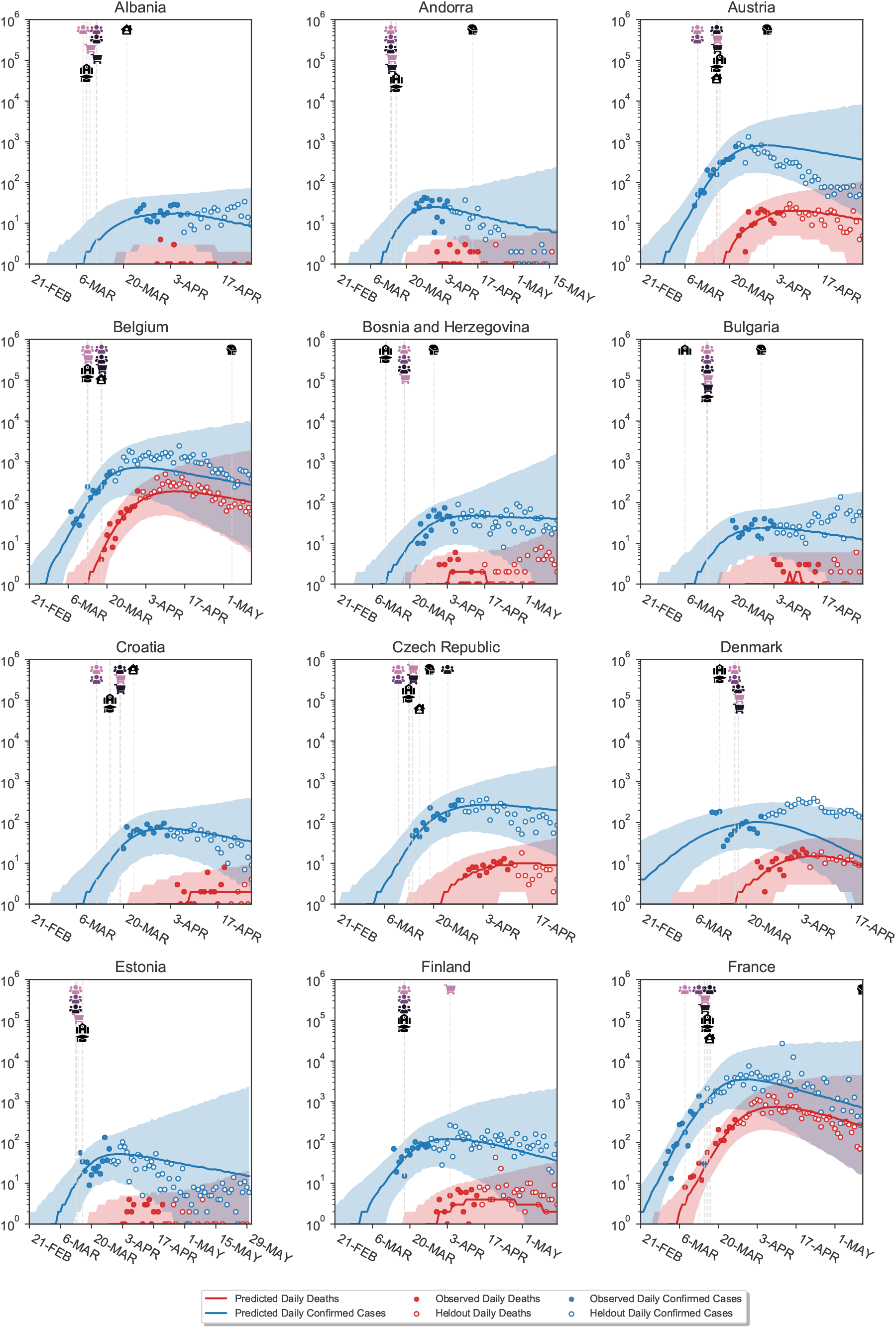
Predictions on excluded countries.

**Figure C.19:**
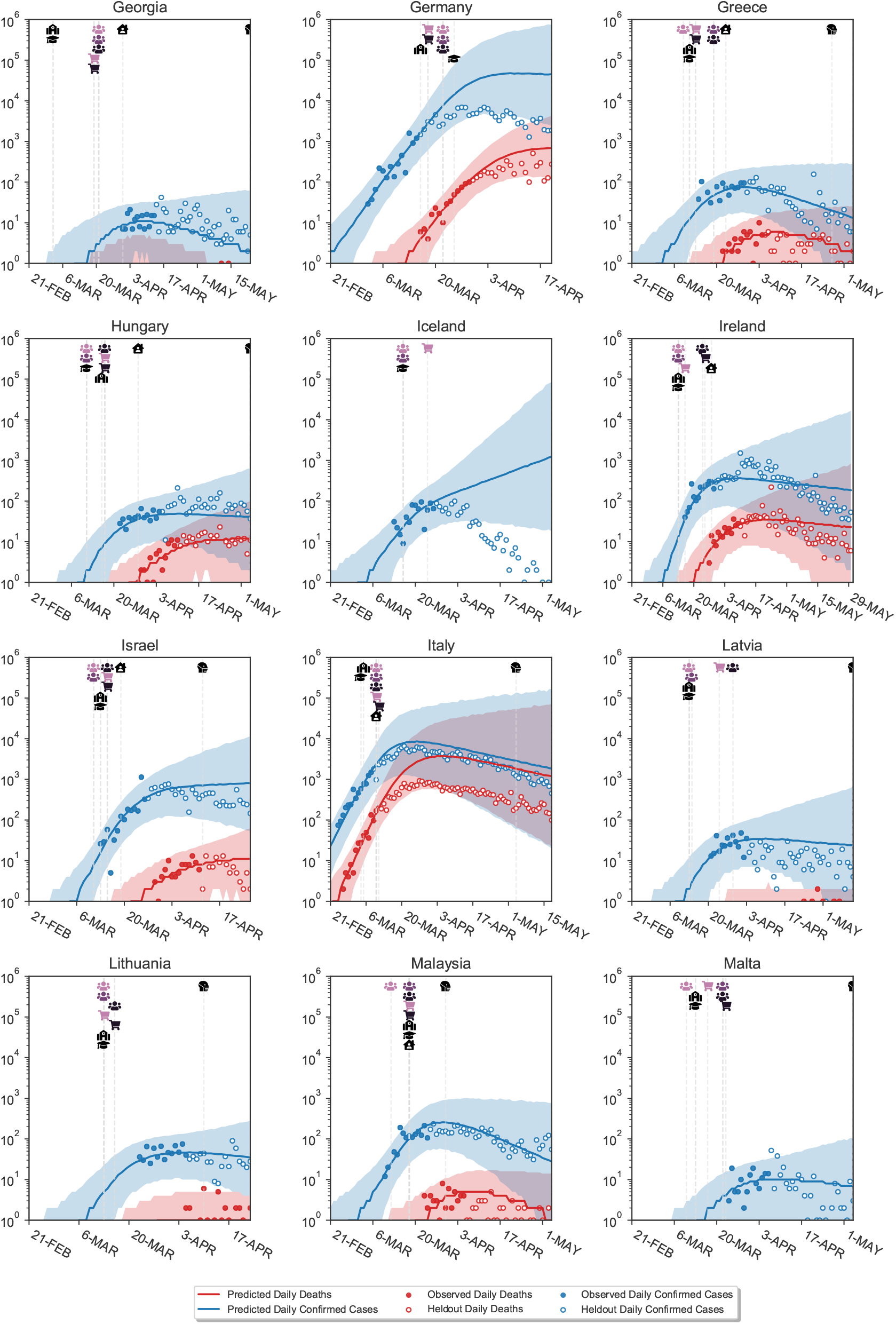
Predictions on excluded countries.

**Figure C.20:**
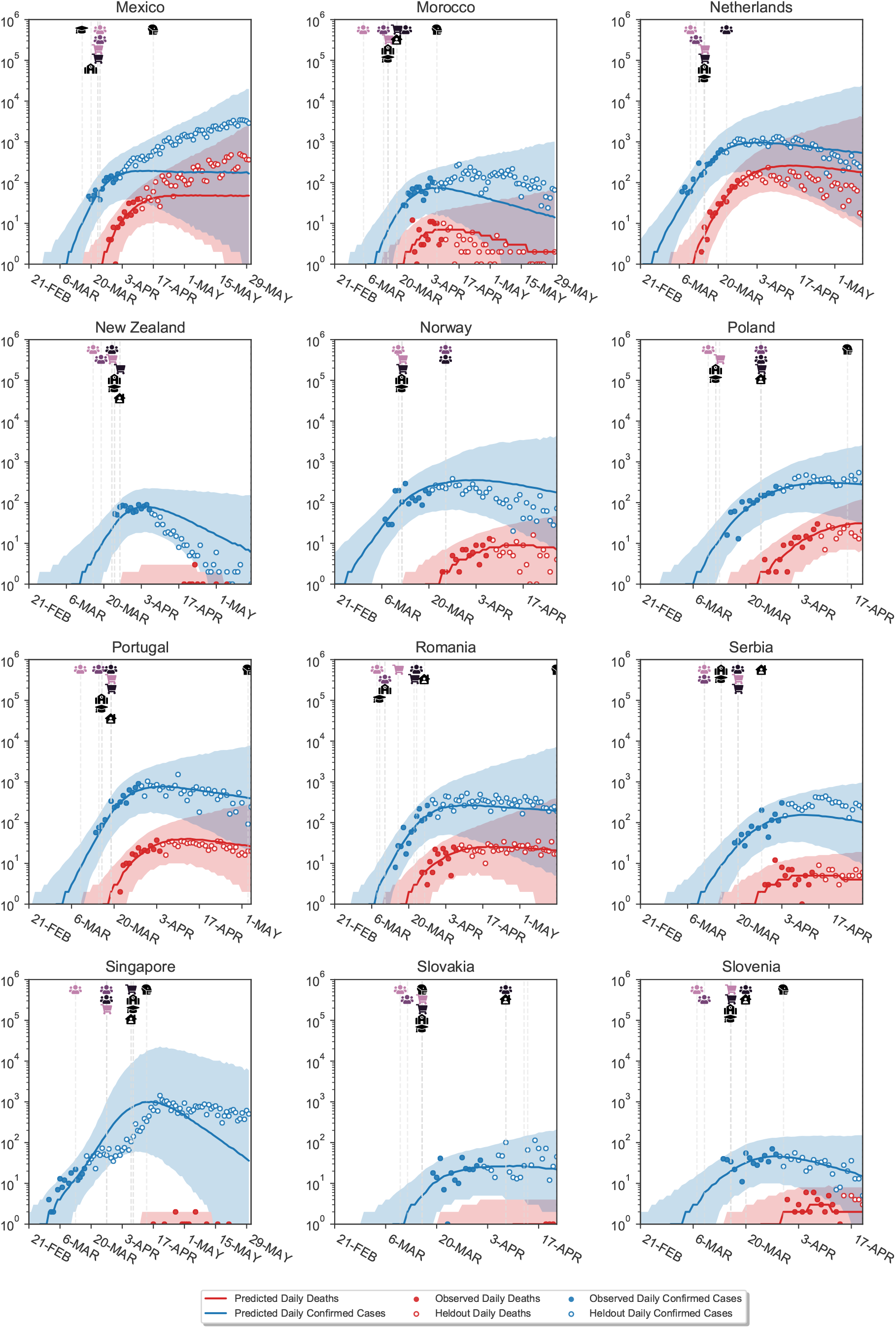
Predictions on excluded countries.

**Figure C.21:**
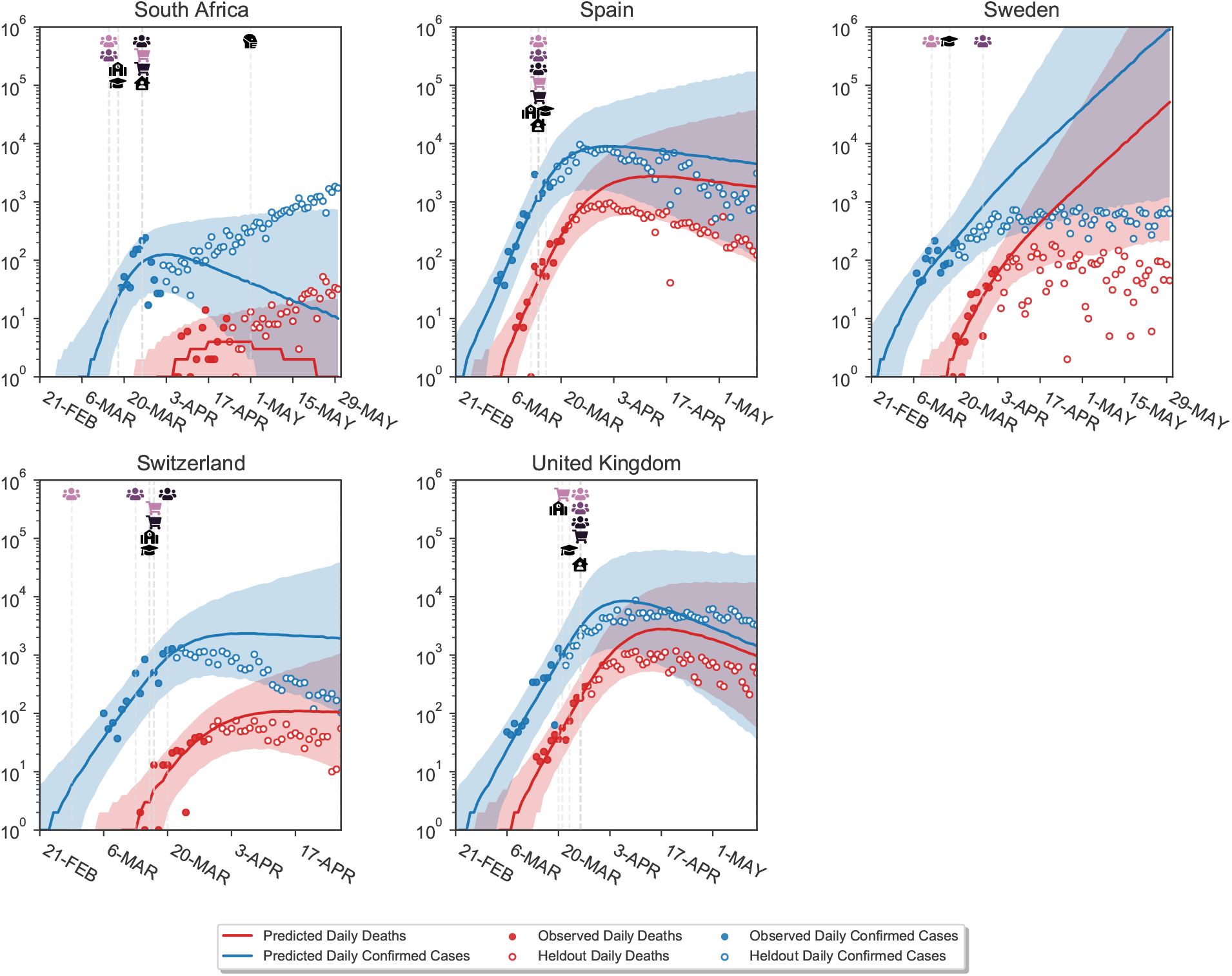
Predictions on excluded countries. Vertical lines show the activation (or inactivation) dates of NPIs. Shaded areas are 95% credible intervals. Blue and red dots show the observed confirmed cases and deaths, while blue and red lines show the median model estimates of cases (*C*_*t*_) and deaths (*D*_*t*_). Empty dots are not observed by the model. For each country, we show the full window of analysis (from the start of the epidemic until the first NPI was lifted, or the 30th of May 2020, whichever was earlier; see Methods).

#### Appendix C.5. Posterior predictive distributions

The posterior predictive distribution (Figure C.22) shows the predicted number of cases and deaths after observing the data. Although these curves can be called ‘fits’, the degree of fit to the data must be interpreted with great care. The fit is generally tight, but this is partly due to the inferred latent noise variables 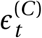 and 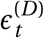. Inferring this latent noise allows the posterior predictive distribution to closely match the data *without overfitting the effectiveness parameters* to the data. Such behavior is common in Bayesian models, which often perfectly interpolate the data without overfitting^33^. The noise terms can account for periods where infections grew faster or slower than predicted based solely on the active NPIs. In such periods, the noise may account for changes in testing, reporting, and unobserved interventions.

**Figure C.22:**
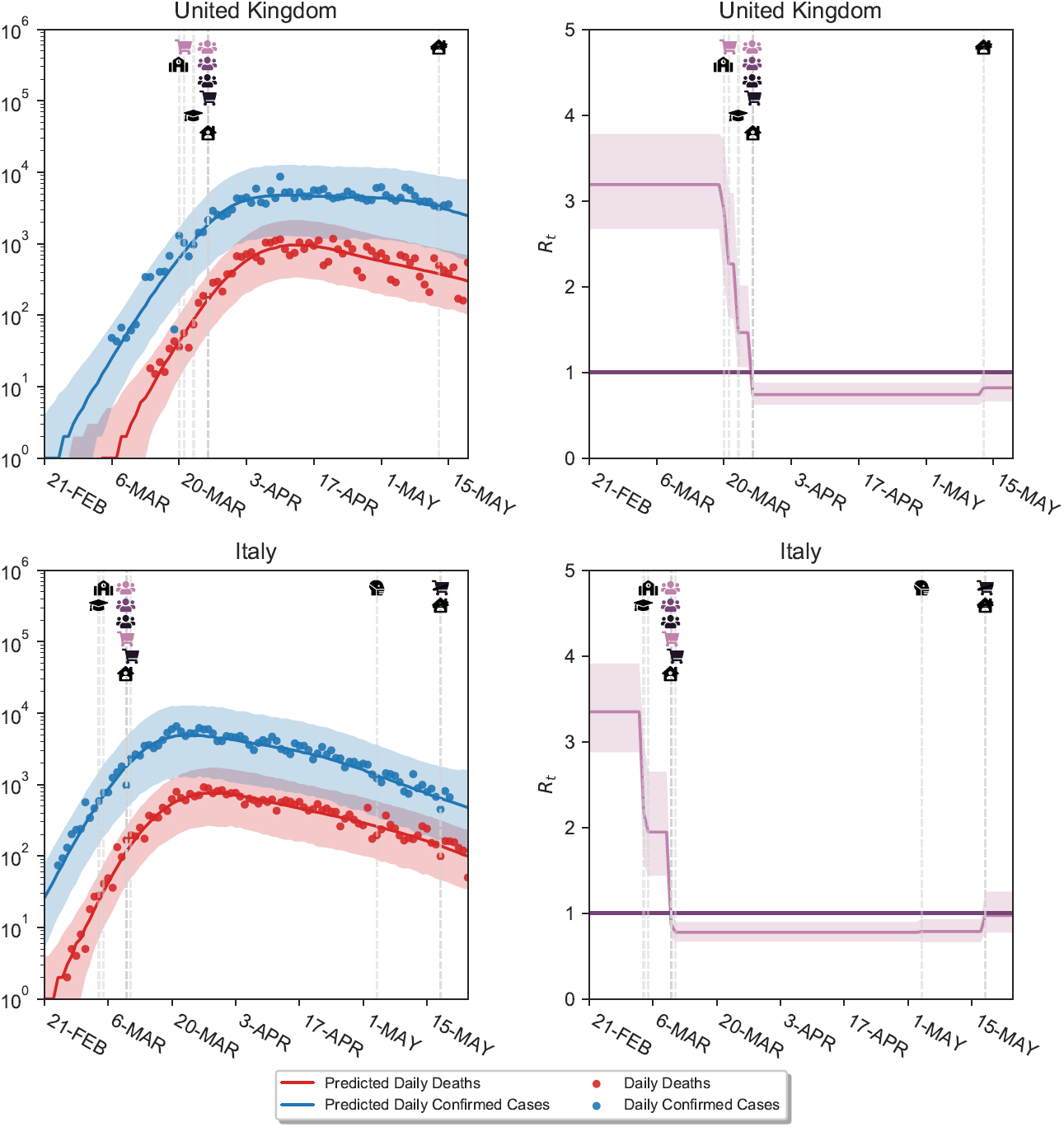
*Left:* Posterior predictive distributions for two exemplary countries. See text. *Right:* Inferred *R*_*t*_ over time.

#### Appendix C.6. Calibration without countries used for hyperparameter selection

**Figure C.23:**
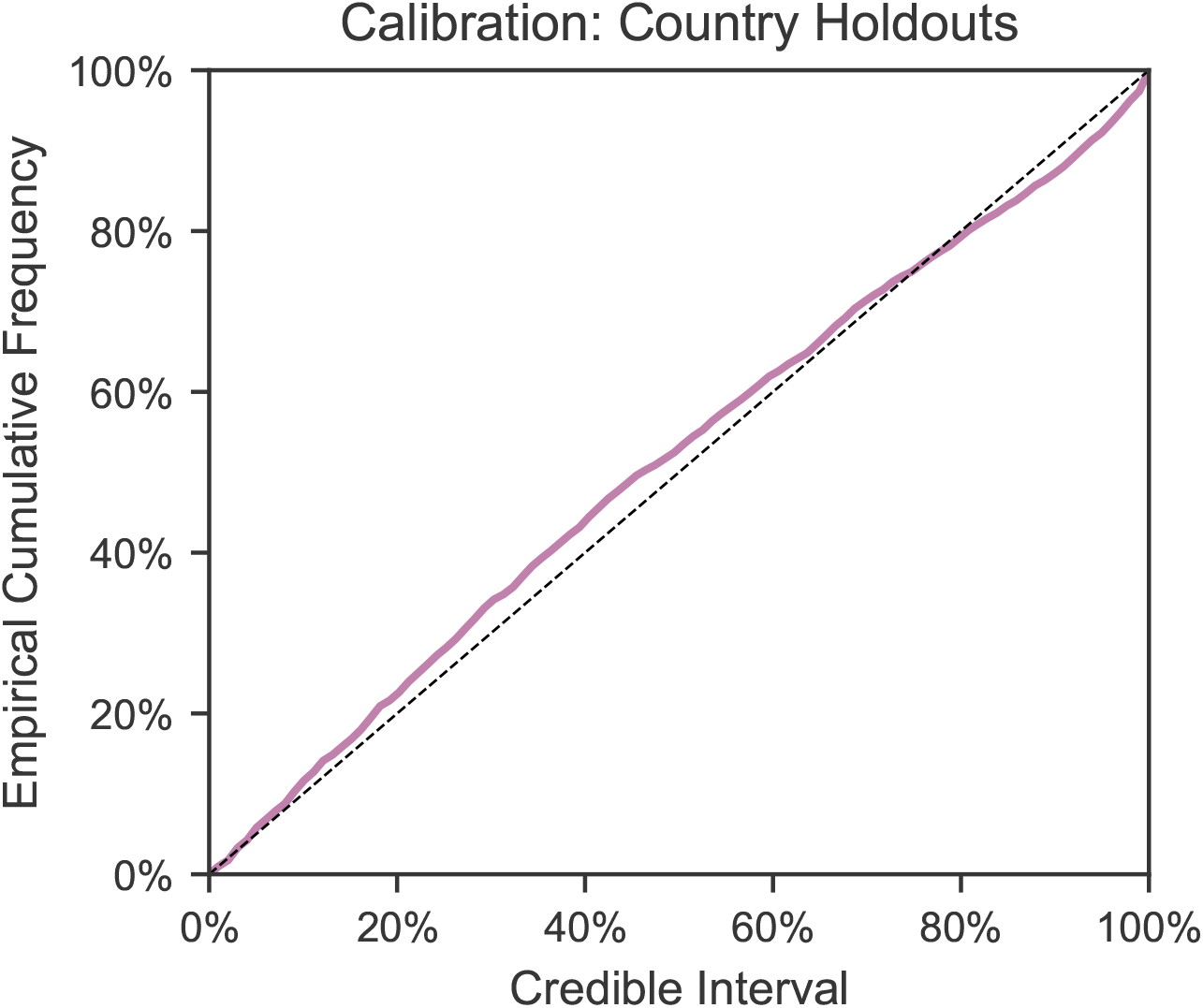
Calibration in held-out countries with leave-one-out cross-validation. Here we include only those 35 countries that were not used to select the hyperparameter. We show the first 14 days of cases and deaths in each country and then extrapolate to future days within the period of analysis. The plot shows the percentage of observed daily case and death counts that lie within the X% credible interval, across all countries and days.

#### Appendix C.7. MCMC stability results

**Figure C.24:**
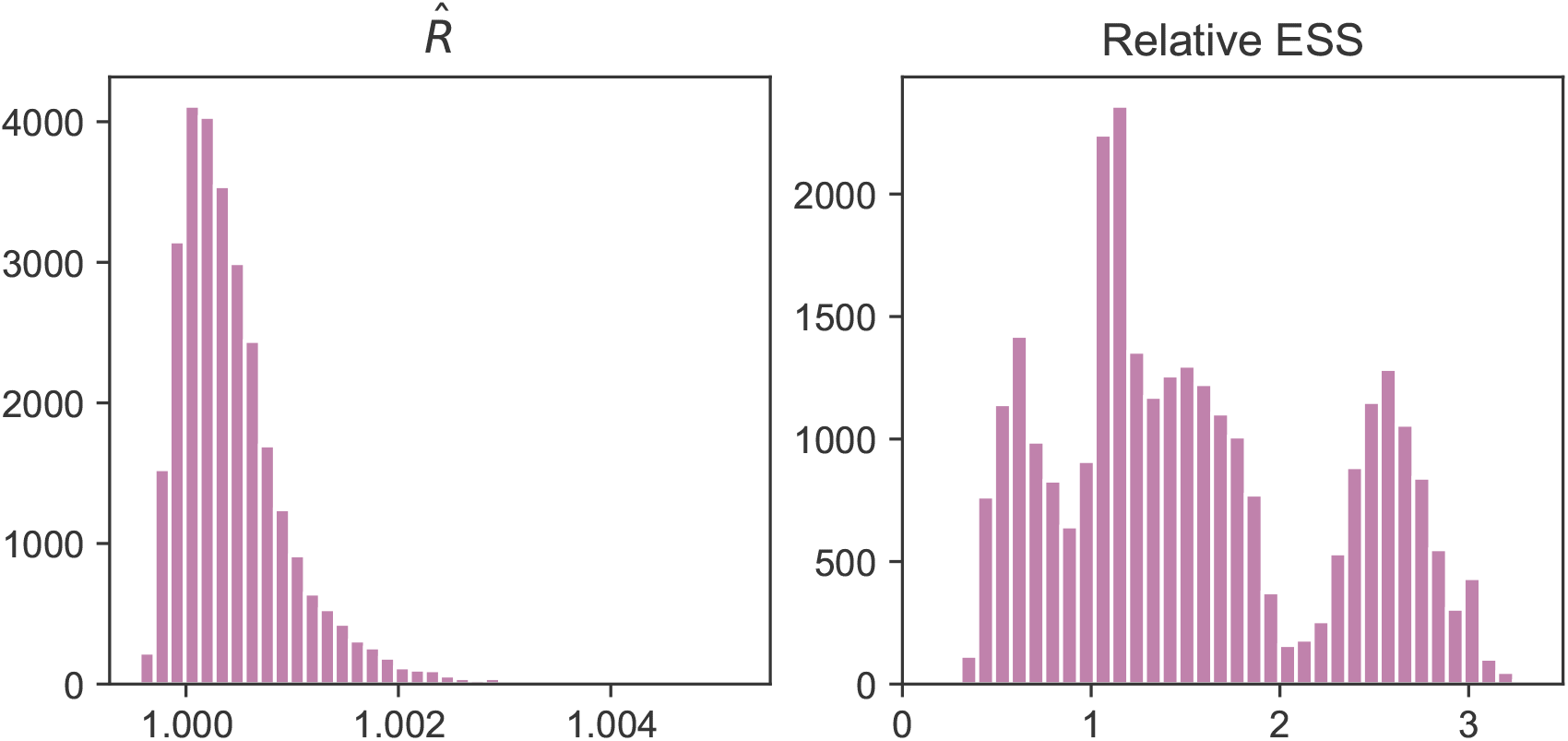
MCMC stability results. *Left:* R-hat statistic. Values are close to 1, indicating convergence. *Right:* Relative effective sample size. A value of 1 indicates perfect decorrelation between samples. Values above (below) 1 indicate that the effective number of samples is higher (lower) than the actual number of samples due to negative (positive) correlation, respectively.

### Appendix D. Additional results

#### Appendix D.1. Estimated *R*_0_ by country

**Table D.5:** Estimated values for *R*_0_ by country. The parenthesis give the 95% credible interval which is often wide. The mean *R*_0_ across countries is 3.3.

| Country | Estimated $R_0$ | Country | Estimated $R_0$ |
| --- | --- | --- | --- |
| Albania | 3.48 (2.75;4.31) | Lithuania | 3.23 (2.48;4.03) |
| Andorra | 2.75 (2.14;3.4) | Malaysia | 2.96 (2.36;3.61) |
| Austria | 3.1 (2.5;3.77) | Malta | 3.27 (2.48;4.14) |
| Belgium | 3.6 (3.02;4.27) | Mexico | 3.99 (3.27;4.8) |
| Bosnia and Herzegovina | 3.31 (2.59;4.06) | Morocco | 3.59 (2.99;4.27) |
| Bulgaria | 3.75 (3.04;4.57) | Netherlands | 3.16 (2.6;3.75) |
| Croatia | 3.42 (2.7;4.18) | New Zealand | 2.41 (1.77;3.1) |
| Czech Republic | 3.46 (2.76;4.25) | Norway | 2.73 (2.15;3.38) |
| Denmark | 2.8 (2.2;3.47) | Poland | 3.97 (3.2;4.82) |
| Estonia | 2.91 (2.29;3.61) | Portugal | 3.55 (2.92;4.25) |
| Finland | 2.79 (2.21;3.43) | Romania | 4.01 (3.35;4.75) |
| France | 3.31 (2.8;3.89) | Serbia | 3.8 (3.08;4.61) |
| Georgia | 3.56 (2.81;4.39) | Singapore | 2.95 (2.38;3.56) |
| Germany | 2.85 (2.31;3.46) | Slovakia | 3.53 (2.71;4.45) |
| Greece | 3.21 (2.61;3.88) | Slovenia | 2.99 (2.3;3.73) |
| Hungary | 4.03 (3.32;4.82) | South Africa | 4.47 (3.77;5.27) |
| Iceland | 1.71 (1.13;2.42) | Spain | 3.6 (3.06;4.23) |
| Ireland | 3.68 (3.09;4.33) | Sweden | 2.29 (1.76;2.88) |
| Israel | 3.79 (3.09;4.59) | Switzerland | 2.89 (2.34;3.51) |
| Italy | 3.36 (2.88;3.91) | United Kingdom | 3.2 (2.67;3.78) |
| Latvia | 3.01 (2.33;3.76) |  |  |

#### Appendix D.2. Collinearity

##### Appendix D.2.1. The individual effects of school and university closures

The dates of school and university closures coincide nearly perfectly for every country except Iceland and Sweden, which closed universities but not schools (Figure 1). As a consequence, the inferred individual effects depend strongly on the inclusion or exclusion of these countries in the dataset (Figure D.25). If we included all countries, we would conclude that university closures were more effective than school closures (black markers in Figure D.25). However, if we excluded Iceland or Sweden, we would conclude that they were roughly equally effective. As there is no strong justification for including or excluding one particular country, we cannot meaningfully disentangle the effects of school and university closures. However, there is much more data to determine the joint effect, and it is indeed much more stable (Figure D.25).

**Figure D.25:**
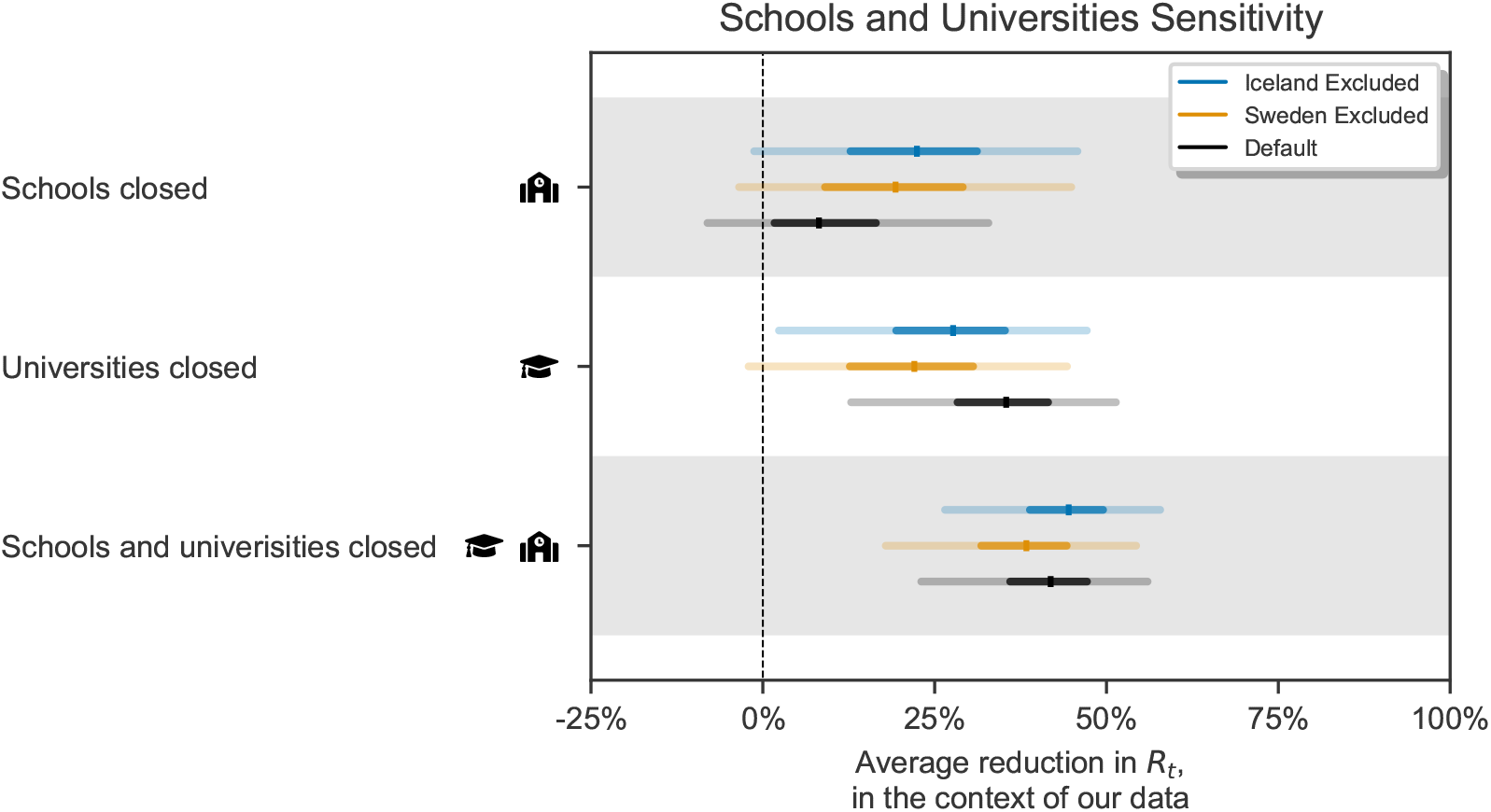
The individual effectiveness of closing schools and of closing universities, as well as the joint effect of closings school *and* universities, estimated on all countries (default), all countries except Sweden, and all countries except Iceland. Median, 50% and 95% credible intervals are shown.

##### Appendix D.2.2. Co-occurrence of NPIs

Table D.6 shows the total number of days across all countries available to distinguish NPI effects. For every pair of NPIs (row - column), the entry shows the number of country-days on which only one of the NPIs was implemented (but not both or neither). Note that we do not show the traditional collinearity statistics (variance inflation factors and data correlations) since their applicability to time series data is limited. In particular, the value of these statistics in our data increases as data for a longer time period becomes available, which would misleadingly suggest that we could address problems from collinearity by using less data.

**Table D.6:** Total number of days across all countries available to distinguish NPI effects. For every pair of NPIs (row column), the entry shows the number of country-days on which exactly one of the NPIs was implemented. Abbreviations: G.: Gatherings; SBC: Some businesses closed; MBC: Most nonessential businesses closed; SaUC: Schools and universities closed; SaHO: Stay-at-home order.

|  | G. <1000 | G. <100 | G. <10 | SBC | MBC | SaUC | SaHO |
| --- | --- | --- | --- | --- | --- | --- | --- |
| Mask-wearing | 1829 | 1686 | 1472 | 1554 | 1315 | 1588 | 1038 |
| Gatherings <1000 |  | 143 | 501 | 299 | 620 | 325 | 1173 |
| Gatherings <100 |  |  | 358 | 240 | 515 | 284 | 1030 |
| Gatherings <10 |  |  |  | 262 | 403 | 308 | 696 |
| Some businesses closed |  |  |  |  | 331 | 176 | 898 |
| Most businesses closed |  |  |  |  |  | 393 | 569 |
| Schools and universities closed |  |  |  |  |  |  | 940 |

##### Appendix D.2.3. Correlations between effectiveness estimates

The effectiveness parameters *α*_*i*_ are typically negatively correlated with each other for NPIs which are often used together, reflecting uncertainty about which NPI is reducing *R*. Excessive collinearity in the data would result in wide posterior credible intervals with strong correlations^24^, but we find weak posterior correlations between effectiveness estimates. The strongest correlation between any pair of NPIs is − 0.42, between “closing schools and universities” and “closing some businesses” (Figure D.26). The weak correlations are one indicator that collinearity is manageable with our dataset.

###### Effect on NPI combinations

To better understand posterior correlations, we visualize their effect in hosted video files. As we condition on different values for one NPI, we can see that the estimates of other NPIs change only slightly, always staying well within the credible intervals in Figure 3. The significance of posterior correlations is small enough that it is possible to calculate a reasonable approximation to the mean effect of a set of NPIs by simply combining the mean percentage reductions for each individual NPI (e.g. two 50% reductions lead to a 75% reduction). For example, this approximation leads to a joint effect of 75% for all NPIs together, which closely matches the exact mean joint effect of 77%.

Videos are available online [here](https://github.com/epidemics/COVIDNPIs/tree/manuscript/online_supplement/results/videos_posterior_correlation).

**Figure D.26:**
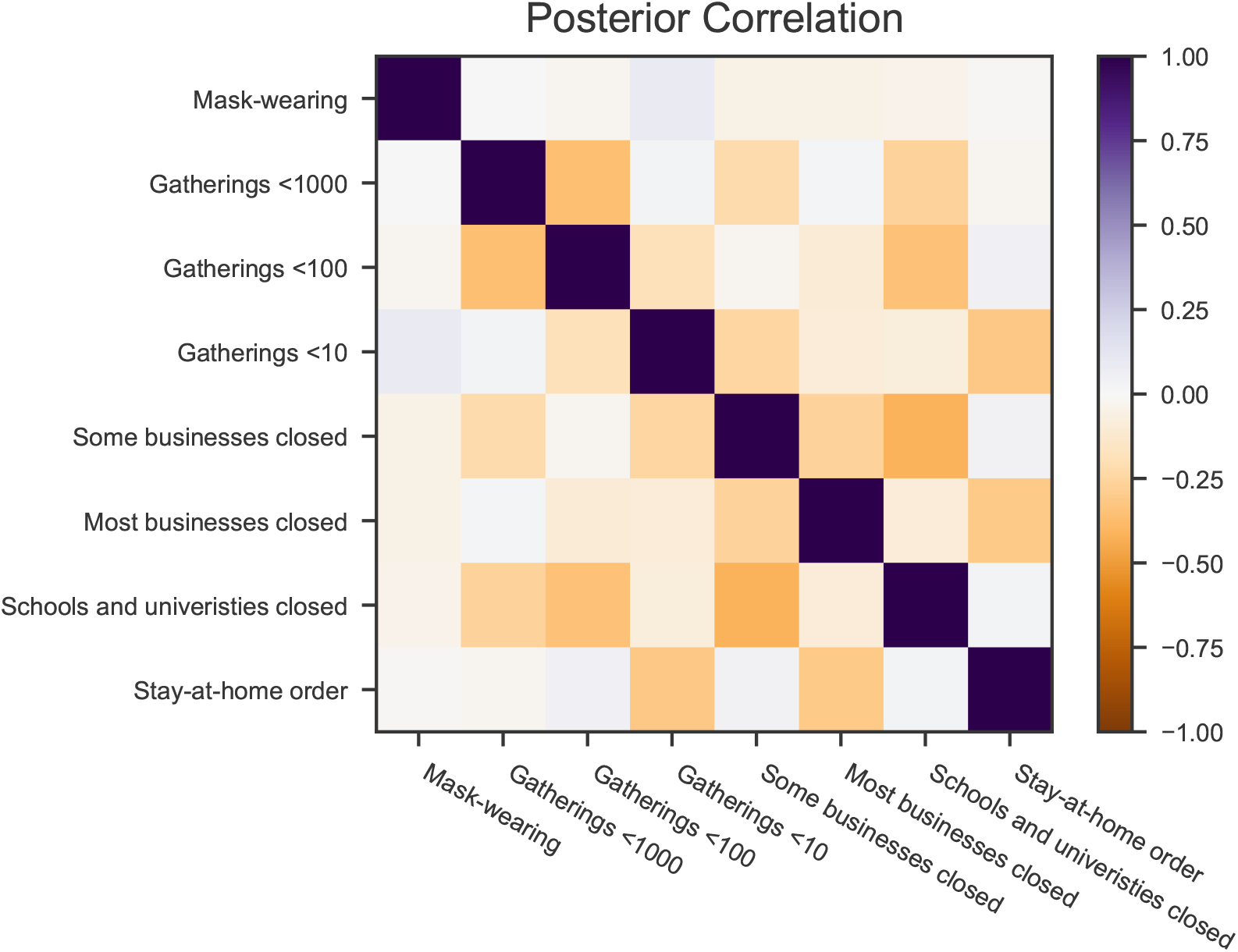
Posterior correlations between effectiveness parameters *α*_*i*_.

#### Appendix D.3. Posterior Epidemiological Parameter Distributions

Figure D.27 shows the posterior distributions for variables describing the key delay distributions in our model: the generation interval, the delay between infection and case confirmation, and the delay between infection and death. We find that the posterior distributions of these parameters are somewhat tighter than their priors, but they still explore a wide range of values. This suggests that the data provides evidence, albeit weak, about these delay distributions (for example, from visual inspection, the data would show that the delay is longer for deaths than for cases). An exception is the dispersion of the infection to death delay distribution, which has a very wide prior.

**Figure D.27:**
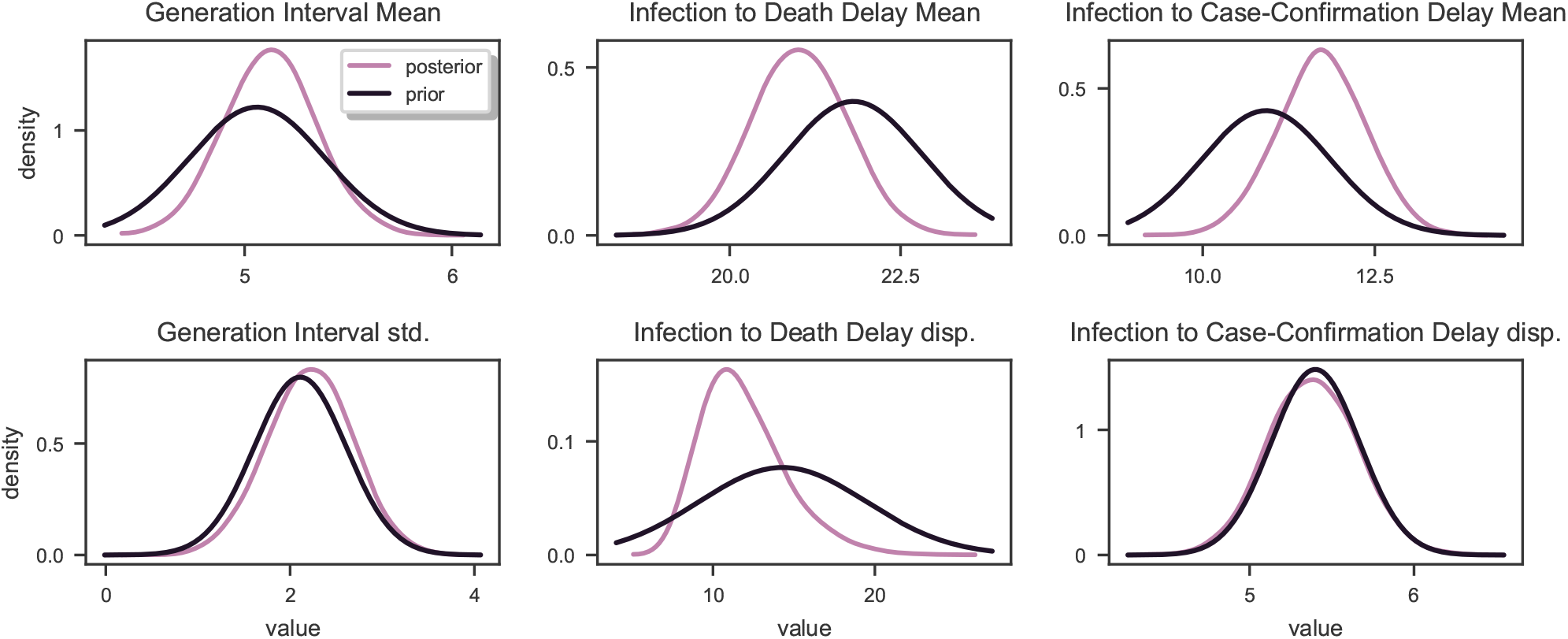
Posterior distributions over key epidemiological parameters describing the generation interval and the delays between infection and case confirmation/death.

### Appendix E. Additional discussion of assumptions and limitations

#### Appendix E.1. Limitations of the data

We only record NPIs if they are implemented in most of a country (if they affect more than three-quarters of the population). We thus miss NPIs which were only implemented regionally. For example, a few regions in Germany implemented stay-at-home orders, but most did not. Thus, Germany is listed as “no stay-at-home order” in our data. Additionally, our NPI definitions were not perfectly granular. For example, a gathering ban on gatherings of >15 people and a ban on gatherings of >60 people would both fall under the NPI “Gatherings limited to 100 people or less”, despite likely having different effects on *R*_*t*_. Finally, while we included more NPIs than previous work (Table F.7), there are many NPIs for which we were not able to collect enough high-quality data for our modeling, such as public cleaning or changes to public transportation.

Of the 41 countries in our dataset, 33 are in Europe. As a result, the NPI effectiveness estimates may be biased towards effects in Europe, and NPI effectiveness may have been different in other parts of the world.

#### Appendix E.2. Model limitations

##### Independence of country and time

We assume that the effect of NPIs on *R*_*t*_ is constant across countries and time. However, the exact implementation and adherence of each NPI is likely to vary. Our uncertainty estimates in Figure 3 account for these problems only to a limited degree. Additionally, different countries have different cultural norms and age profiles, affecting the degree to which a particular intervention is effective. For example, a country where a higher proportion of the population is in education will likely experience a larger effect from a government order to close schools and universities. Our estimates thus should be adjusted to local circumstances. To address differences between countries, our structural sensitivity analysis includes a model where each NPI can have a different effect per country (Appendix B.2). The average effectiveness estimates across countries in this model match the conclusions from our default model.

##### Testing, reporting, and the IFR

Our model can account for differences in testing (and IFR/reporting) between countries and over time, as discussed in Appendix A. However, we have not used additional data on testing to validate if it does so reliably. Our model may struggle to account for changes in the testing regime—for instance, when a country reaches its testing capacity so that the ascertainment rate declines exponentially. An exponential decline would have the same effect on observations as an unobserved NPI. Consequently, we cannot quantify its effect on our results (though the sensitivity analyses look reassuring).

##### Interaction between NPIs

As discussed in the Results section, our model reports the average additional effect each NPI had in the contexts where it was active in our data (in the sense mathematically shown by Sharma et al.^9^). Figure 3 (bottom left) summarises these contexts, aiding interpretation. The effectiveness of an NPI can only be extrapolated to other contexts if its effect does not depend on the context. For example, we may expect that closing schools has a similar effectiveness whether or not businesses are also closed. But wearing masks in public may be less effective when a stay-at-home order limits public interactions.

##### Growth rates

The functional form of the relationship between the daily growth rate of the number of infections *g*_*t*_ and the reproductive number *R*_*t*_ holds exactly when the epidemic is in its exponential growth phase, but becomes less accurate as the number of susceptible people in a population decreases and/or control measures are implemented. However, we also report results from a *renewal process* model^8^ that does not depend on this assumption, and we find similar effectiveness estimates.

##### Signalling effect of NPIs

As we explained in the Discussion for school closures, we do not distinguish between the direct effect of an NPI and its indirect effect as it signals the gravity of the situation to the public. Conversely, lifting interventions may also have a signalling effect.

##### Homogeneous effect of interventions

We work under the implicit assumption that NPIs affect different population groups equally. This could affect results in various ways. For example, suppose country A tests an older demographic than country B, and we are considering the effect of an NPI that mostly affects the older demographic (for example, isolating the elderly). Then the NPI will appear to have a greater effect on confirmed cases in country A, breaking the assumption that effects are constant across countries. Our previous discussion of interpreting results when this assumption is violated applies.

### Appendix F. Overview of previous work

**Table F.7:** Data-driven, multi-country, multi-NPI studies of the effectiveness of observed (as opposed to hypothetical) NPIs in reducing the transmission of COVID-19.

| Study | NPIs studied | Regions/countries studied | Method |
| --- | --- | --- | --- |
| Banholzer et al., 2020<sup>23</sup> | School closure, border closure, event ban, gathering ban, venue closure, lockdown, work ban | U.S., Canada, Australia, Austria, Belgium, Denmark, Finland, France, Germany, Greece, Ireland, Italy, Luxembourg, the Netherlands, Portugal, Spain, Sweden, UK, Norway, Switzerland | Semi-mechanistic Bayesian hierarchical model |
| Chen and Qiu, 2020<sup>21</sup> | Travel restriction, mask-wearing, lockdown, social distancing, school closure, centralized quarantine | Italy, Spain, Germany, France, UK, Singapore, South Korea, China, U.S. | Regression with delayed effect Susceptible-Infectious-Removed (SIR) model |
| Flaxman et al., 2020<sup>1</sup> | School or university closure, case-based isolation, ban on large public events, social distancing, lockdown | Austria, Belgium, Denmark, France, Germany, Italy, Norway, Spain, Sweden, Switzerland, UK | Semi-mechanistic Bayesian hierarchical model |
| Islam et al., 2020<sup>20</sup> | School closures, workplace closures, restrictions on mass gatherings, public transport closure, lockdown | 149 countries or regions | Interrupted time series regression |
| Liu et al., 2020<sup>22</sup> | 13 NPIs from the OXCGRT dataset | 130 countries and regions | panel regression |

**Table F.8:** Some data-driven studies of the effectiveness of observed NPIs in reducing the transmission of COVID-19, which study a single-country and/or a single-NPI.

| Study | NPIs studied | Regions/countries studied | Method |
| --- | --- | --- | --- |
| Choma et al., 2020<sup>34</sup> | Single aggregated NPI | 22 countries and 25 states | Regression with Susceptible-Infectious-Removed-Deceased (SIRD) model |
| Dandekar and Barbastathis, 2020<sup>35</sup> | General quarantine and isolation | Wuhan, Italy, South Korea, and U.S. | A mix of a mechanistic model and a data-driven neural network model |
| Dehning et al., 2020<sup>36</sup> | Contact ban, restrictions on gatherings, schools, childcare, businesses | Germany | Bayesian inference of transmission rate |
| Gatto et al., 2020<sup>37</sup> | Various restrictions to mobility and human-to-human interactions | Italy | Susceptible–Exposed–Infected–Recovered (SEIR)-like disease transmission model |
| Hsiang et al., 2020<sup>19</sup> | Restricting travel (5 subcategories), distancing (10 subcategories), quarantine and lockdown (2 subcategories), additional policies (2 subcategories) | China, South Korea, Italy, Iran, France, U.S. (each country modelled individually) | Linear regression on estimated growth rates |
| Jarvis et al., 2020<sup>38</sup> | Physical (social) distancing measures | UK | Questionnaire data and compartmental epidemic model |
| Kraemer et al., 2020<sup>39</sup> | Travel restrictions and cordon sanitaire | China | Regression |
| Kucharski et al., 2020<sup>40</sup> | Travel restrictions | Wuhan (China) | Various, including Susceptible-Exposed-Infectious-Removed (SEIR) model |
| Lai et al., 2020<sup>41</sup> | Case detection and isolation, travel restrictions, contact reductions | China | Travel-network-based SEIR model |

Table F.8 – *Continued from previous page*
| **Study** | **NPIs studied** | **Regions/countries studied** | **Method** |
| --- | --- | --- | --- |
| Lorch et al., 2020<sup>42</sup> | Mobility restrictions, testing & tracing, social distancing, and business restrictions | Tübingen (Germany) | Authors' own spatiotemporal model of epidemics |
| Maier and Brockmann, 2020<sup>43</sup> | General quarantine and isolation | Mainland China | Quantitative fits to empirical data |
| Orea and Álvarez, 2020<sup>44</sup> | Lockdown | Spain | Spatial econometric analysis |
| Quilty et al., 2020<sup>45</sup> | Intercity travel restrictions | Beijing, Chongqing, Hangzhou, and Shenzhen (Mainland China) | Branching process transmission model |
| Sears et al., 2020<sup>46</sup> | Mobility changes as a proxy for stay-at-home mandates | U.S. | Difference-in-differences statistical model |
| Siedner et al., 2020<sup>47</sup> | General social distancing | U.S. | Interrupted time-series |

### Appendix G. Handling edge cases in the data collection

In our data collection process, we relied on carefully worded definitions of 9 different NPIs (Table F.7), which allowed us to systematically determine the date on which a country imposed an NPI and, if applicable, the date the NPI was lifted.

In some cases, however, we faced ambiguities in how to interpret the start date of an NPI. One kind of challenge arose when descriptions of policy measures were less specific than our NPI definitions (e.g. a ban on “large gatherings” that does not specify the exact number of people that constitutes a “large gathering”). Another difficulty was due to NPI policies that made distinctions that we did not make in our own NPI definitions (e.g., an NPI policy that made a distinction between the number of people able to gather indoors vs outdoors).

To resolve these ambiguities in a consistent manner, our researchers developed a set of principles and guidelines that were followed during the data collection process. For each of the examples below, the relevant sources are available in the data table in the supplementary material.

#### Situation: Sometimes only public gatherings are banned, with no explicit ban on private gatherings

*How we deal with it:* We still counted this as a ban on gatherings.

Examples:

- Sweden: In Sweden, they banned all *public* gatherings of more than 50 people (demonstrations, religious meetings, theater performances, markets, and other events that relied on the constitutional freedom of assembly), however, the ban did not have a mandate to prohibit *private* gatherings (such as private parties). We counted this as a ban on gatherings.
- Finland: In Finland, they banned all public gatherings of more than 10 people on the 16th of March. Although formal restrictions did not apply to private gatherings, this policy met our definition of a ban on gatherings. (Note that this inclusion seems particularly valid in light of the fact that, according to Finnish police, the formal restrictions on public events were widely interpreted to apply to private gatherings as well, and there were very few reports of large private parties despite the absence of formal restrictions.)

#### Situation: The size limits on gatherings sometimes differ between indoor and outdoor gatherings

*How we deal with it:* In these cases, we relied on the limitations on indoor events, as these events entail a greater risk of transmission.

Example:

- Spain: In Spain, a range of rules were employed as the country gradually eased restrictions on gatherings. In phase 1, cultural events were permitted with up to 30 people indoors and up to 200 outdoors. This was counted as “Gatherings limited to 100 people or less.”

#### Situation: The size limit on gatherings sometimes differs between different types of gatherings

*How we deal with it:* In this case, researchers would use their best judgment to infer whether the restriction would apply to *most* gatherings of a given size.

Example:

- Spain: In Spain, phase 1 of the reopening allowed for cultural events to have up to 30 participants indoors, while social gatherings were limited to 10 people. In this case, since “cultural events” is broad, we counted this as a case of “gatherings limited to 100 people or less.” However, if for example all gatherings above 5 people had been banned with an exception for funerals, we would have counted this as “gatherings limited to 10 people or less,” since the exemption only applied to a minority of gatherings.

#### Situation: Limitations on gathering sizes are not clearly given, yet a policy stating that “large events are banned” is in place

*How we deal with it:* Our researchers used the relevant context to infer the most likely scope of the policy.

Example:

- Albania: on March 8 “authorities had also ordered cancellations of all large public gatherings including cultural events and were asking sporting federations to cancel scheduled matches”. The events that are mentioned here are multi-thousand person gatherings, and so we took March 8th to be the start date of “Gatherings limited to 1000 people or less”. However it was unclear whether gatherings of 100-1000 would also have been banned, so we did not yet say that “Gatherings limited to 100 people or less” was instantiated.

#### Situation: Only some schools were closed, or schools reopened gradually

*How we deal with it*: Since our definition of the NPI is that “*Most* schools are closed”, we did not count the closure of just a few schools or school years sufficient to meet this criteria. Similarly, if schools reopened for only a very limited number of year groups, for example for final year students sitting exams, we did not count this as a lifting of the “most schools closed” NPI.

Examples:

- Sweden: Sweden kept all schools through 9th grade open, but closed high schools (>16 year olds). In this case, we did not count this as “Most schools closed”, since more than 75% of students are below 9th grade.
- Czech Republic: After closing all schools on March 13, the Czech Republic allowed schools to reopen for teaching in some contexts from May 11 (specifically for students in their final year of primary school or high school preparing for exams). However, we still counted this as “Most schools closed” since the majority of students were not in school. We recorded the end date for school closure to be June 8, when all schools reopened.

#### Situation: In a country where most non-essential businesses were closed, the lifting of business closures is gradual, and businesses in different sectors are successively allowed to open

*How we deal with it:* Countries reopen sectors in different, idiosyncratic ways and successions. Given the available data, it is not feasible to create a principle that can be applied unambiguously to every single case without some involvement of researcher judgment. The general guideline we used was: If only a few, low-risk businesses (e.g., bike stores, hardware stores, etc.) are additionally allowed to reopen, then we still counted this as “Most nonessential businesses closed.” However if any *one* of the following criteria are met, then we counted “Most nonessential businesses closed” as having lifted, but the “Some businesses closed” NPI was still in place:

- All regular retail stores, with only a few exceptions e.g. size limitations, are open
- Contact-based services, such as hairdressers and tattoo parlors, are open
- Restaurants and bars are open and serving indoors

We decided that meeting any one of these criteria is a sufficient condition for taking a country from “Most nonessential businesses closed” to “Some businesses closed.” This heuristic was partly based on the fact that the status of these categories appeared to be consistently correlated, meaning that, even in the absence of complete specifications as to what had reopened or not, it was typically possible to infer the overall level of reopening based on either of these categories. Meeting at least one of these criteria was considered a necessary condition for ending the “Most nonessential businesses closed” NPI.

Examples:

- Slovakia: On April 22, retail operations and services up to 300 *m*^2^ opened. Since this meets one of the sufficient conditions, we counted April 22 as the end date for “Most nonessential businesses closed”
- Ireland: On May 18, the following reopened: hardware stores, builders, merchants and those providing essential supplies, retailers involved in the sale and repair of vehicles, certain office supply stores. Because this white list does not meet any of the three criteria, Ireland’s end date for “Most nonessential businesses closed” was not counted as May 18.
- Czech Republic: On April 20, several businesses reopened, including farmer’s markets, marketplaces, locksmiths, bike shops, car dealers, electronics stores. At this point, none of the criteria were met, so we recorded the Czech Republic as still having “Most nonessential businesses closed”. On May 11, a long list of businesses reopened, including barbers, hairdressers, museums, all establishments in sufficiently large shopping centers, shows with up to 100 participants, and restaurants with a window facing the street. Since contact-based services (hairdressers) and all retail establishments in sufficiently large spaces were allowed to reopen, we counted May 11 as the end date for the “Most nonessential businesses closed” NPI.
- Croatia: On April 27, all “trade activities” (except within shopping malls), service jobs that don’t involve physical contact, museums, libraries, and galleries opened. Since the criteria regarding “all retail stores being open” was met, we counted April 27 as the end date for “Most nonessential businesses closed”.

### Appendix H. All model equations

This section will mainly be of interest to readers that wish to re-implement the model. Variables are indexed by NPI *i*, country *c*, and day *t*. All prior distributions are independent.

#### Data

1. **NPI Activations**: *ϕ*_*i,t,c*_ ∈ {0, 1}.
2. **Observed (Daily) Cases: Ct**,**c**.
3. **Observed (Daily) Deaths**: *D*_*t,c*_.

#### Prior Distributions

1. **Country-specific** *R*_0_: *R*_0,*c*_ ∼ Normal(3.25, *κ*); *κ* ∼ Half Normal(*µ =* 0, *σ =* 0.5).
2. **NPI effectiveness:** *α*_*i*_ ∼ `Asymmetric Laplace`(*m =* 0, *κ =* 0.5, *λ =* 10). *m* is the location parameter, *κ >*0 is the asymmetry parameter, and *λ >*0 is the scale parameter.
3. **Infection Initial Counts:**

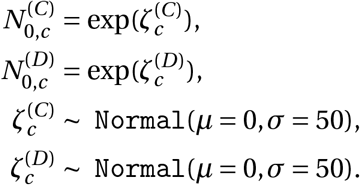
4. **Observation Noise Dispersion Parameters:**

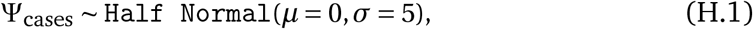

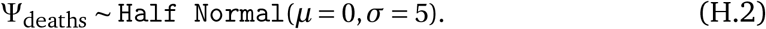

#### Hyperparameters

1. **Growth Noise Scale**, *σ*_*g*_ *=* 0.2.

#### Delay Distributions

1. **Generation interval distribution**^13,14^:

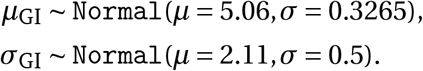
2. **Time from infection to case confirmation** 𝒯 ^(*C*)13,15,16^:^f^

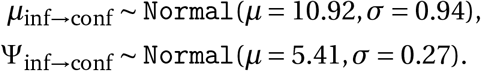 This distribution is converted into a forward-delay vector:

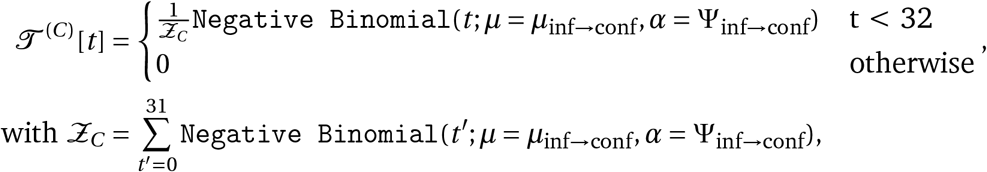

i.e., the delay follows a truncated and normalised negative binomial distribution.
3. **Time from infection to death** 𝒯 ^(*D*)**f**13,15,17^:

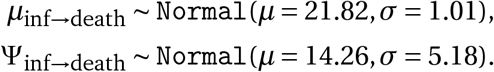 This distribution is converted into a forward-delay vector:

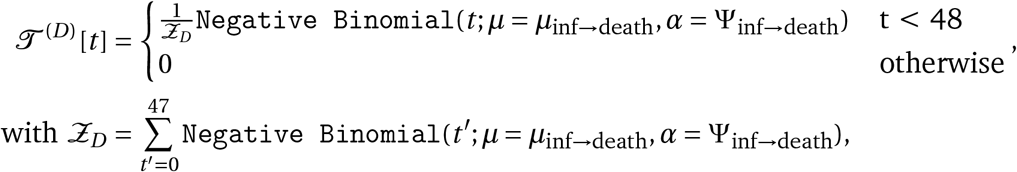

i.e., the delay follows a truncated and normalised negative binomial distribution.

#### Infection Model

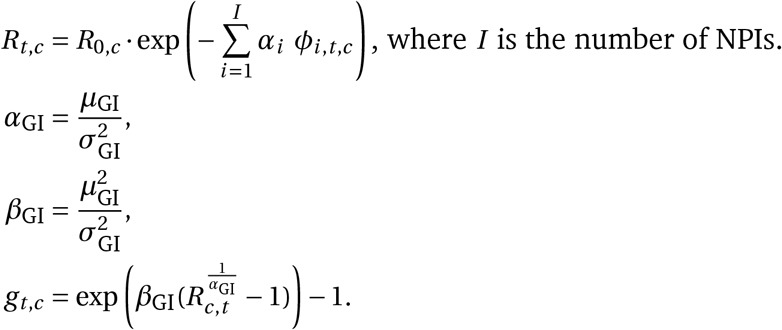

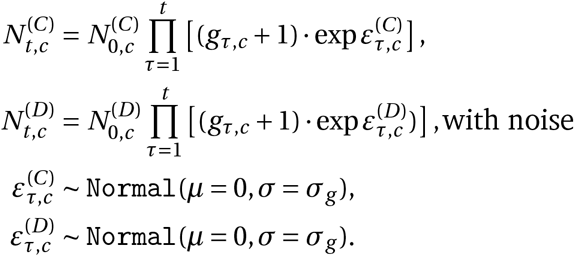

#### Observation Model^f^

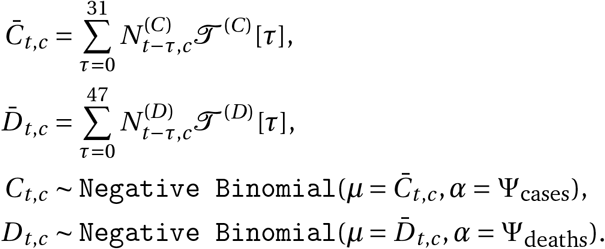

## Footnotes

a The countries were selected for the availability of reliable NPI data at the time when we started data collection and modelling (April 2020); and for their presence in at least one of the public datasets that we used to cross-validate our collected data. We excluded countries with fewer than 100 cases (or 10 deaths) by March 31, as our model neglects new cases and deaths below these thresholds. We also excluded a small number of countries if there were credible media reports casting doubt on the trustworthiness of their reporting of cases and deaths. Finally, we excluded very large countries like China, the US, and Canada, for ease of data collection, as these would require more locally fine-grained data. 33 of the 41 included countries are in Europe. As a result, the NPI effectiveness estimates may be biased towards effects in Europe, and NPI effectiveness may have been different in other parts of the world.

b Concretely, the window of analysis extended until three days after the first reopening for confirmed cases, and 13 days after the first reopening for deaths. These values correspond to the 5% quantile of the infection-to-confirmation/death distributions, ensuring that less than 5% of the new infections on the reopening day were still observed in the window of analysis.

c We evaluated the following datasets: - Epidemic Forecasting Global NPI Database^14^ - Oxford COVID-19 Government Response Tracker (OxCGRT)^15^ - ACAPS #COVID19 Government Measures Dataset Note that these datasets are under continuous development. Many of the mistakes found will already have been corrected. We know from our own experience that data collection can be very challenging. We have the fullest respect for the people behind these datasets. In this paper, we focus on a more limited set of countries and NPIs than these datasets contain, allowing us to ensure higher data quality in this subset. Given our experience with public datasets and our data collection, we encourage fellow COVID-19 researchers to independently verify the quality of public data they use, if feasible.

d In principle, a stay-at-home order does not imply these other NPIs. It is possible for a country to simultaneously have allowed schools to be open and have a stay-at-home order (with exemptions for attending school) in place. However, this is not how stay-at-home orders were implemented by the countries in our dataset, and we cannot estimate the effect of such a hypothetical stay-at-home order from the available data.

a Many epidemiological models define growth rates as the exponent *r* in an exponential growth function. Here, we use daily growth rates instead for ease of exposition. These choices are mathematically equivalent. Note that we adapted equation (2.9) in Wallinga & Lipsitch^5^ to account for our choice.

b Since we treat new infections as a continuous number, its initial value can be between 0 and 1.

c This is only approximately true. The negative binomial output distribution has a coefficient of variation diminishing with its mean; i.e., smaller observations are relatively more noisy and carry less weight. Furthermore, whilst the prior over 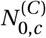 could break scale invariance, the uninformative prior results in a negligible effect.

d However, our model may struggle when the ascertainment rate also changes exponentially over time. This could happen when a country reaches its testing capacity. See Appendix E.

e The main qualitative conclusions as outlined in the Discussion are: Stay-at-home orders had a small effect; Mandating mask-wearing had a small effect; School and university closures had a large effect; Gatherings bans were effective; More strict gathering bans were more effective than less strict ones; Business closures were effective; Closing most nonessential businesses had limited benefit over closing just high-risk businesses.

f *α* in the definition of the Negative Binomial distribution is the dispersion parameter. Larger values of *α* correspond to a *smaller* variance, and less dispersion. With our parameterisation, the variance of the Negative Binomial distribution is 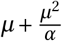.

